# WOMEN’S PRECONCEPTION HEALTH, PLANNING, AND BEHAVIOURS: A CROSS-SECTIONAL SURVEY OF PREGNANT WOMEN IN AUSTRALIA

**DOI:** 10.64898/2026.07.10.26357787

**Authors:** Amie Steel, Jenny Hall, Adina Lang, Erica McIntyre, Jon Adams, Wendy Burton, Danielle Schoenaker

**Affiliations:** School of Public Health, Faculty of Health, University of Technology Sydney, Ultimo NSW Australia; Reproductive Health Research Department, EGA Institute for Women’s Health, UCL, London, UK; Synergistiq, 552 Victoria Street, North Melbourne, Melbourne, VIC, 3051, Australia; Institute for Sustainable Futures, University of Technology Sydney, Ultimo NSW Australia; Research Institute for Innovative Solutions for Well-being and Health (INSIGHT), University of Technology Sydney, Ultimo NSW Australia; Royal Australian College of General Practitioners, Antenatal and Postnatal Specific Interest Group, Brisbane QLD Australia; School of Human Development and Health, Faculty of Medicine, University of Southampton, Southampton, UK; MRC Lifecourse Epidemiology Centre, University of Southampton, Southampton, UK; NIHR Southampton Biomedical Research Centre, University of Southampton and University Hospital Southampton NHS Foundation Trust, Southampton, UK

**Keywords:** preconception, pregnancy, health behaviours, women’s health, folic acid, social determinants of health

## Abstract

**Aim:** To determine the prevalence, degree and timing of pregnancy planning among pregnant women in Australia and identify the specific health behaviours with which pregnant women engage during specified preconception timeframes.

**Methods:** A national retrospective cross-sectional survey conducted December 2020-September 2021 including a convenience sample of pregnant women aged 18-49-years in Australia and their reproductive partners, recruited through social media platforms.

**Results:** Overall, 615 eligible women with relevant survey items were included in this study with 59.6% (95% CI 55.6-63.4) of pregnancies categorised as ‘planned’, 38.3% (95% CI 34.5–42.2) as ‘ambivalent’ and 2.1% (95% CI 1.2–3.6) as ‘unplanned’. Unplanned/ambivalent pregnancies were most common in women under 20 years old and least common in women aged 30 to 39 years (p=0.05). They were also most common among women who were married (56.6% vs 47.8%; p=0.001), university qualified (70.3% vs 52.6%; p<0.001) and in full-time employment (56.9% vs 44.9%, p=0.002). Actions to improve preconception health were generally uncommon; however, they were more likely in women who planned their pregnancies. For example, 42.7% of women with ambivalent/unplanned pregnancies reported consuming folic acid/pregnancy multivitamin before pregnancy compared with 83.3% among those who planned their pregnancy (p<0.001). Preconception financial status, BMI and general health were also associated with pregnancy planning (p<0.05).

**Conclusion:** Almost all women in this survey identified some level of pregnancy planning – including ‘planned’ and ‘ambivalent’ pregnancies - yet actions to improve their preconception health were uncommon. In addition to promoting specific actions that individuals can undertake with regards to preconception health, it is vital that the impact of structural barriers and wider determinants of health (e.g., out-of-pocket health costs, health literacy, and clear identification of health professionals trained to deliver preconception care) be adequately considered and addressed in terms of preconception health promotion, planning and policy. Such a broad approach can help strengthen attempts to improve pregnancy planning and preconception health.

## INTRODUCTION

Optimising one’s health prior to conception (preconception) is increasingly recognised in the literature and health systems as critical to improving a couple’s chances of successfully becoming pregnant and having a healthy pregnancy and child [1, 2]. Evidence has grown over the past two decades highlighting the crucial influence of parental health behaviours and wellbeing around the time of conception on the environment in which eggs and sperm mature [3]. These factors can affect epigenetic changes to the programming and development of the embryo impacting the health of the newborn child and across the lifespan, known as the developmental origins of health and disease (DOHaD) [3-5]. Poor maternal preconception health (PCH) can increase the risk of pregnancy complications such as gestational diabetes and hypertension [6, 7], and affect the long-term cardiometabolic disease risks for mothers and their children [8-10]. In addition to these cardiometabolic comorbidities, maternal obesity can increase the risk of infertility, pregnancy loss and complications during labour and delivery [11]. Parental preconception exposures include modifiable risk factors such as dietary intake and micronutrient supplementation consistent with national guidelines, regularly undertaking physical activity, maintaining a healthy weight, smoking, alcohol and recreational drug cessation, chronic disease management, health screening and relevant up-to-date immunisations [12, 13].

While the body of PCH research continues to grow, there remains limited adoption of targeted, routine preconception care (PCC) practices among health professionals to promote and support the prevention of chronic diseases and optimisation of health before pregnancy [12]. PCC is an approach that includes recommending medical and behavioural interventions to women and their partners before conception [2]. The Royal Australian College of General Practitioners (RACGP) recommend screening for pregnancy intention, and provision of advice on relevant medical risk factors, lifestyle, immunisation, and folic acid and iodine supplementation to women planning to conceive [13]. Chronic conditions pose a significant burden on individuals, communities and health systems, accounting for the largest number of deaths internationally [14] and in Australia [15]. The Australian Government’s National Strategic Framework for Chronic Conditions has responded with an emphasis on prevention and health promotion from conception across the life course [16].

The preconception period represents an ideal time for couples to receive health advice and engage in counselling to optimise their health and wellbeing and reduce these risks in preparation for pregnancy [1]. Despite the available guidelines and growing public health interest in PCC,[17] only a few isolated PCC service providers currently exist in Australia [18] and internationally [19]. Furthermore, the literature has shown low awareness and knowledge of the benefits of PCH behaviour change [20]. Global estimates in high-income countries suggest 20% to 80% of pregnancies are intended, with wide variation depending on country, socioeconomic status and type of measure used [1, 21]. Previous studies have explored and measured the prevalence of pregnancy planning and associated preconception behaviours among pregnant women in Australia [12, 22, 23]. However, these studies were limited by recruitment in antenatal care settings and there is no nationwide data describing pregnancy planning among women in Australia using a validated measure. A clear understanding of the prevalence of pregnancy planning and women’s health behaviours in the preconception period is needed to enable the development of responsive strategies, including increasing the uptake of PCC services.

This study aims to directly address this need and provide a more comprehensive picture of the state of pregnancy planning in Australia. To achieve this, the current study recruited women and their reproductive partners more broadly via social media, giving an opportunity to include women who were not planning to continue their pregnancy to participate in the study and allowing recruitment of women across Australia.

## METHODS

### Aims

This study aimed to determine the prevalence, degree and timing of pregnancy planning among pregnant women in Australia. It also aimed to identify which health behaviours pregnant women engaged in during specified preconception timeframes.

### Study design and setting

This observational cross-sectional retrospective survey study involved a convenience sample of pregnant women in Australia and their reproductive partners. It was conducted between December 2020 and September 2021 using an online survey instrument.

### Participant eligibility and recruitment

Participants were eligible to participate if they self-identified as a woman aged between 18 and 49 years living in Australia and at any stage of pregnancy, or a reproductive partner – defined as the male who provided the sperm that contributed to the pregnancy - of someone that met these criteria. They were recruited using a targeted advertisement on social media platforms (i.e., Twitter, Facebook, Instagram). Participants from each sample group (i.e., pregnant women and reproductive partners) were incentivized to participate by offering a random prize draw of $100 to individuals who completed the survey. The analysis presented in this paper only includes the participants who completed the survey for pregnant women (i.e., not for reproductive partners).

### Survey administration and data management

All data were collected using the Qualtrics^TM^ online survey platform. Interested parties accessed the screening instrument via a link on the social media advert. Individuals who completed the screening instrument were sent an email containing the link for the survey targeting their group (e.g., pregnant women) and a second survey with a link for the survey targeting the second group (e.g., reproductive partners),) which they were asked to forward to their respective partner via email.

### Survey instrument

The 92-item survey instrument (see Supplementary File 1 – Survey Instrument) was developed with reference to questionnaires first developed and used in the United Kingdom [24]. It incorporated the London Measure of Unplanned Pregnancy (LMUP) which had been validated in the Australian population [25]. The full survey was also pilot-tested for face and functional validity by three individuals who matched the eligibility criteria of the target sample population. Minor refinements to wording and survey flow were made based on their feedback. The instrument collected information across five domains: 1) participant demographics (16 items), 2) description of current pregnancy (15 items), 3) preconception and pregnancy health behaviours (38 items), 4) preconception and pregnancy health information and advice (13 items), and 5) health history and general health status (10 items). The analysis presented in this paper uses selected survey items relevant to the research question, covering participant demographics, current pregnancy status, preconception health behaviours, and general health status.

### Statistical analysis

Data were imported into Stata 17.1 for analysis. The study view rate was determined by the number of individuals who accessed the screening instrument and provided their email address. The survey participation rate was calculated based on the number of people who completed the first survey item directly related to preconception health behaviour (“How long before becoming pregnant were you thinking about having a baby?”), divided by the individuals who completed the screening instrument and provided their email address. The completion rate was calculated as the number of participants completing the last survey item divided by the number who participated in the study.

Participants were analysed for representativeness to the national population by testing the state of residence and age of participants against the Australian women who gave birth in 2021, as described in the Australia’s Mothers and Babies report [26]. Differences in state and age between the sample and target population were tested using chi-square tests.

London Measure of Unplanned Pregnancy (LMUP) scores were calculated consistent with the instrument’s guide [27]. Due to a technical issue with the online survey design, the final item of the LMUP, which should have permitted participants to select multiple items, only permitted one item to be selected. For this reason, participants could only be awarded a maximum score of ‘1’ for this item, instead of the usual maximum of ‘2’. A new variable was then generated that allocated respondents to one of three categories in line with their LMUP score: ‘Planned’ (10 or more), ‘Ambivalent (between 4 and 9) or ‘Unplanned’ (3 or less). Prevalence estimates were calculated to identify the confidence interval associated with the frequencies for each category. A binary LMUP variable was then generated which combined the ‘Ambivalent’ and ‘Unplanned’ categories (i.e., ‘Planned’ and ‘Ambivalent/Unplanned’).

Missing data for the LMUP items were handled in accordance with the LMUP scoring guide, whereby participants who provided response to less than three items were excluded from the analysis using case-wise deletion while participants who missed one to three items had values imputed. Once the cases had been identified, any remaining missing data from other survey items were excluded from the different tests as appropriate to the requirements of the test.

Body mass index (BMI) was calculated using participants reported height and weight prior to pregnancy. New variables were then generated to group these data according to the World Health Organization classifications [28]. Data were reported descriptively with categorical variables presented as frequencies and percentages and continuous variables presented as medians with interquartile ranges. Chi-square (categorical variables), Wilcoxon-ranked (normative continuous) or ANOVA (non-normative continuous) tests were employed to compare differences in participants’ characteristics based on their allocated LMUP categories, using the binary LMUP variable (‘Planned’ and ‘Ambivalent/Unplanned’). Shapiro-wilks tests were used to determine normative distribution.. <u>The effect size of associations between BMI classification and preconception health behaviours pertaining to physical activity or exercise, and to following a weight loss diet were calculated using Cramer’s V analysis. The Cramer’s V scores were then categorised according to a 6-point scale as ‘negligible’ (0.00 to <0.10), ‘weak’ (0.10 to <0.20), ‘moderate’ (0.20 to <0.40), ‘relatively strong’ (0.40 to <0.60), ‘strong’ (0.60 to<0.80) or ‘very strong association’ (0.80 to <1.00) [29].</u> Women’s preconception care behaviours were compared with the Royal Australian College of General Practitioner Guidelines for Preventive Activities Prior to Pregnancy [13].

The most parsimonious model of variables describing the characteristics of women with planned pregnancies compared with women with unplanned/ambivalent pregnancies was determined using staged backwards logistic regression. In accordance with an epidemiological understanding of social determinants of health, a hierarchical conceptual framework [30] was applied to the regression through which the variables were grouped as: demographics, health history, and preconception health behaviours. All variables from each group with an alpha value of less than 0.2 resulting from their binary comparisons were included in the analysis. The regression models were undertaken in stages based on this hierarchy and in the order outlined above. Once the backwards elimination was completed for the first group (demographics), the variables for the next group (health history) in the hierarchy were added and the backwards logistic regression analysis repeated. A similar process was followed for the final group (preconception health behaviours), after which a final model was achieved. A Firth correction was applied to the regression analysis to account for non-convergence outputs resulting from small sample bias detected for key variables. Significance was defined as an alpha value of ≤0.05.

### Ethical conduct of research

This study was conducted in accordance with the principles of the Declaration of Helsinki. The study was evaluated and approved by the University of Technology Sydney Medical and Health Research Ethics committee (#ETHI21-6461). Informed consent was obtained from all participants.

## RESULTS

The study screening instrument was accessed and completed by 908 respondents, of whom 615 (67.7%) were eligible to be included in the analysis. Participants’ LMUP scores identified 59.6% (95% CI 55.6-63.4) of pregnancies as ‘planned’, 38.3% (95% CI 34.5 – 42.2) as ‘ambivalent’ and 2.1% (95% CI 1.2 – 3.6) ‘unplanned’ (see Supplementary File 2 – London Measure of Unplanned Pregnancy Item Response and Scores).

### Demographics

Table 1 presents the demographic characteristics of participants. Most participants were in New South Wales (31.2%), Victoria (22.6%) and Queensland (22.8%), and were aged between 20 years and 29 years (45.8%) or 30 years and 39 years (52.6%). This differed from the national population by both state of residence (p=0.004) and age (p<0.001). A statistically significant difference in participant demographics was identified between women with ‘planned’ and ‘ambivalent/unplanned’ pregnancies. ‘Ambivalent/unplanned’ pregnancies were most common in women under 20 years old and least common in women 30 to 39 years old (p=0.05). Women with ‘planned’ pregnancies more commonly married (56.6% vs 47.8%; p=0.001) or with a university qualification (70.3% vs 52.6%; p<0.001) or employed full time (56.9% vs 44.9%, p=0.002). A greater proportion of women with ‘planned’ pregnancies reported their financial status as ‘not too bad’ (48.9% vs 42.1%) or ‘easy’ (30.2% vs 19.4%) compared to women with ‘ambivalent/unplanned’ pregnancies (p<0.001). Women with ‘planned’ pregnancies also had a higher rate of private health insurance with obstetrics cover (30.2% vs 21.9%, p=0.05) and a lower rate of having a health care card (20.3% vs 27.5%).

**Table 1:**
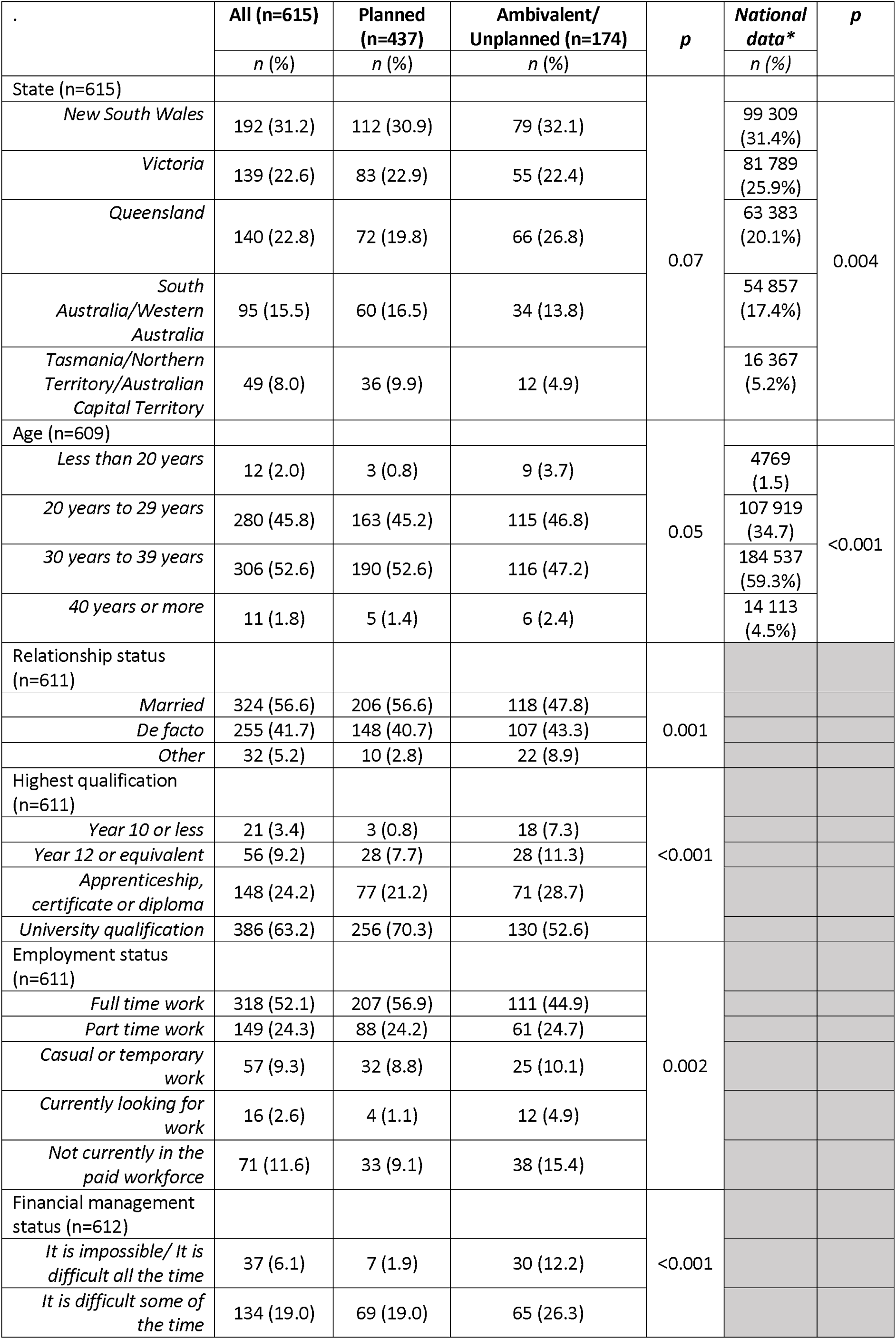
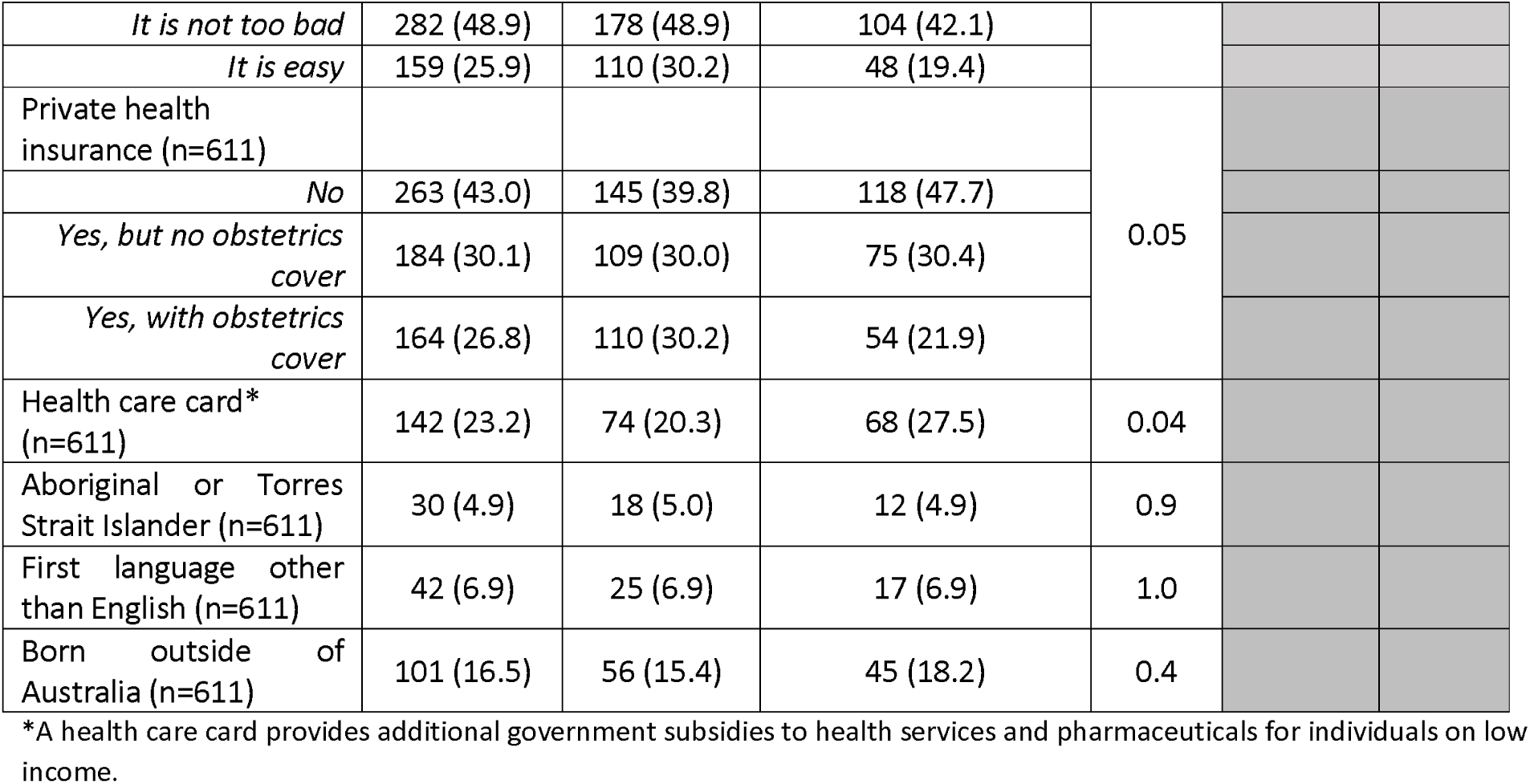
Participant demographic characteristics.

### Health history and general health status

There was a significant association between general health status and LMUP category (Table 2), with a greater proportion of individuals reporting excellent health among those with ‘planned’ pregnancy compared to ‘ambivalent/unplanned’ pregnancy (22.7% vs 11.5%; p<0.001). A greater proportion of women with ‘ambivalent/unplanned’ pregnancy reported having a mental health, psychiatric or neurological condition (33.7%) compared to women with a ‘planned’ pregnancy (25.0%; p=0.05). The mean BMI among participants was 26.7 kg/m^2^, with a significantly lower median BMI among women with ‘planned’ pregnancy (BMI 24.7) compared to women with ‘ambivalent/unplanned’ (BMI 25.7) (p=0.04). A lower proportion of women with a ‘planned’ pregnancy reported having ever smoked (30.0%) compared to those who had an ‘ambivalent/unplanned’ pregnancy (38.0%; p=0.05). There was a significant difference between the groups (p<0.001) in the use of contraception in the six months prior to pregnancy; individuals with an ‘ambivalent/unplanned’ pregnancy more commonly reported not using a contraceptive and not trying to conceive (36.9%) compared to women with a ‘planned’ pregnancy (14.2%). Participants reported a median of one previous pregnancy. Less than one third (30.7%) of participants reported previously given birth at least once. This rate was higher among women who reported an ‘ambivalent/unplanned’ pregnancy (37.9%; p=0.004).

**Table 2:**
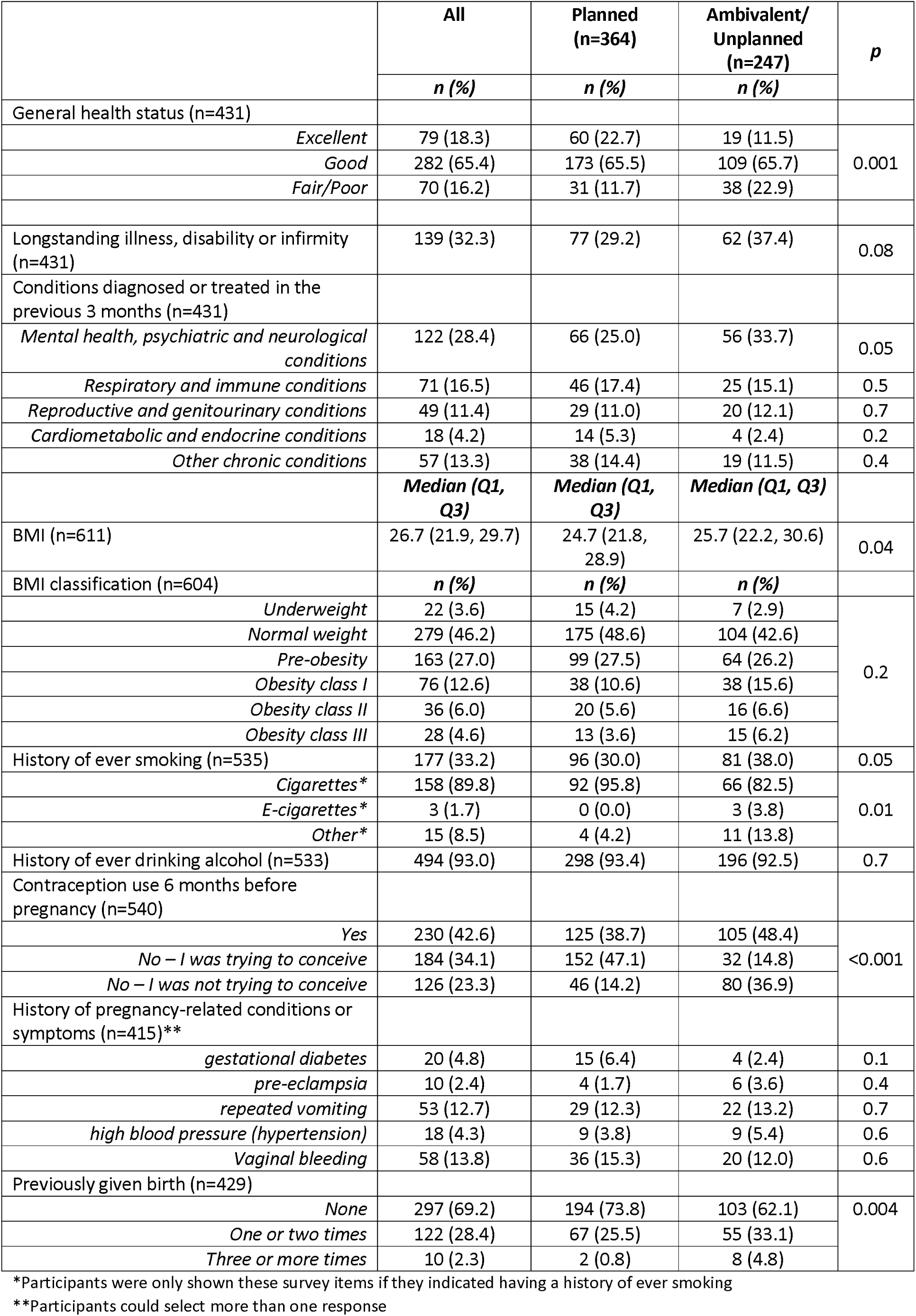
Participant health history and general health status.

### Pregnancy characteristics

Participants had a median of 23 weeks gestation at survey completion and 4 weeks gestation at pregnancy confirmation (see Table 3). ‘Planned’ pregnancy was associated with earlier pregnancy confirmation (p<0.001). Most commonly, participants confirmed their pregnancy using a home pregnancy test (93.8%). A small proportion of participants (8.2%) became pregnant using fertility treatments. Over half of participants reported attempting pregnancy for more than three months (3-5 months: 15.2%; 6-12 months: 17.0%; >12 months: 26.2%). This trend was significantly associated with participants’ LMUP category (p<0.001); it was more pronounced among participants with a ‘planned’ pregnancy (3-5 months: 19.2%; 6-12 months: 21.7%; >12 months: 33.5%) compared to other women.

**Table 3:** Participant pregnancy characteristics.

| Pregnancy characteristics | All | Planned<br>(n=364) | Ambivalent/<br>Unplanned (n=247) | *p* |
| --- | --- | --- | --- | --- |
|  | Median (Q1,<br>Q3) | Median (Q1,<br>Q3) | Median (Q1, Q3) |  |
| Weeks of pregnancy at time of survey completion (n=611) | 23 (15, 30) | 23 (16, 30) | 22 (14, 30) | 0.6 |
| Weeks of pregnancy at time of pregnancy confirmation (n=611) | 4 (4, 5) | 4 (4, 5) | 5 (4, 5) | <0.001 |
| Pregnancy confirmation method (n=611) | *n (%)* | *n (%)* | *n (%)* |  |
| *Home pregnancy test* | 573 (93.8) | 347 (95.3) | 226 (91.5) | 0.07 |
| *From a health professional* | 34 (5.6) | 14 (3.9) | 20 (8.1) |  |
| *Other* | 4 (0.7) | 3 (0.8) | 1 (0.4) |  |
| Duration trying for pregnancy before conceiving (n=611) |  |  |  |  |
| *I was not trying to become pregnant* | 113 (18.5) | 3 (0.8) | 110 (41.1) | <0.001 |
| *Less than one month* | 70 (11.5) | 41 (11.3) | 29 (11.7) |  |
| *Between one and two months* | 71 (11.6) | 49 (13.5) | 22 (8.9) |  |
| *Between three and five months* | 93 (15.2) | 70 (19.2) | 23 (9.3) |  |
| *Between six and twelve months* | 104 (17.0) | 79 (21.7) | 25 (10.1) |  |
| *More than twelve months* | 160 (26.2) | 122 (33.5) | 38 (15.4) |  |
| Pregnancy resulting from fertility treatment (n=611) | 50 (8.2) | 38 (10.4) | 12 (4.9) | 0.01 |

### Preconception health behaviours

Table 4 presents the frequency of health behaviours recommended for women during the preconception period. Less than half of participants reported engaging in a range of protective health behaviours prior to becoming pregnant including visiting the dentist (46.5%) or checking immunisations were up to date (36.9%) in the 12 months before conception. Some participants reported following a weight loss diet during the six months before becoming pregnant (20.4%). A much greater proportion (75.7%) reported undertaking physical activity or exercise during the three months before becoming pregnant, with a median of five hours of physical activity per week reported by participants. Further analyses investigated the relationship between following a preconception weight loss diet or engaging in preconception physical activity and participant BMI category (see Table 5). The analysis found a moderate association between the prevalence of weight loss diet adherence in the six months preconception and participant BMI category (V 0.220, p<0.001), whereby the participants categorised as Obesity Class I had the highest proportion of individuals following a weight loss diet (39.1%) while no ‘underweight’ participants indicating following a diet for weight loss. A weak association was also observed between participant physical activity in the three months preconception and BMI category (V 0.194, p=0.001) as more than half of participants in each category reported preconception physical activity except for those categorised as ‘obesity class III’ (46.2%). The highest proportion of participants engaging in preconception physical activity were within the ‘normal weight’ (80.8%) and obesity class II (77.4%) categories.

**Table 4:**
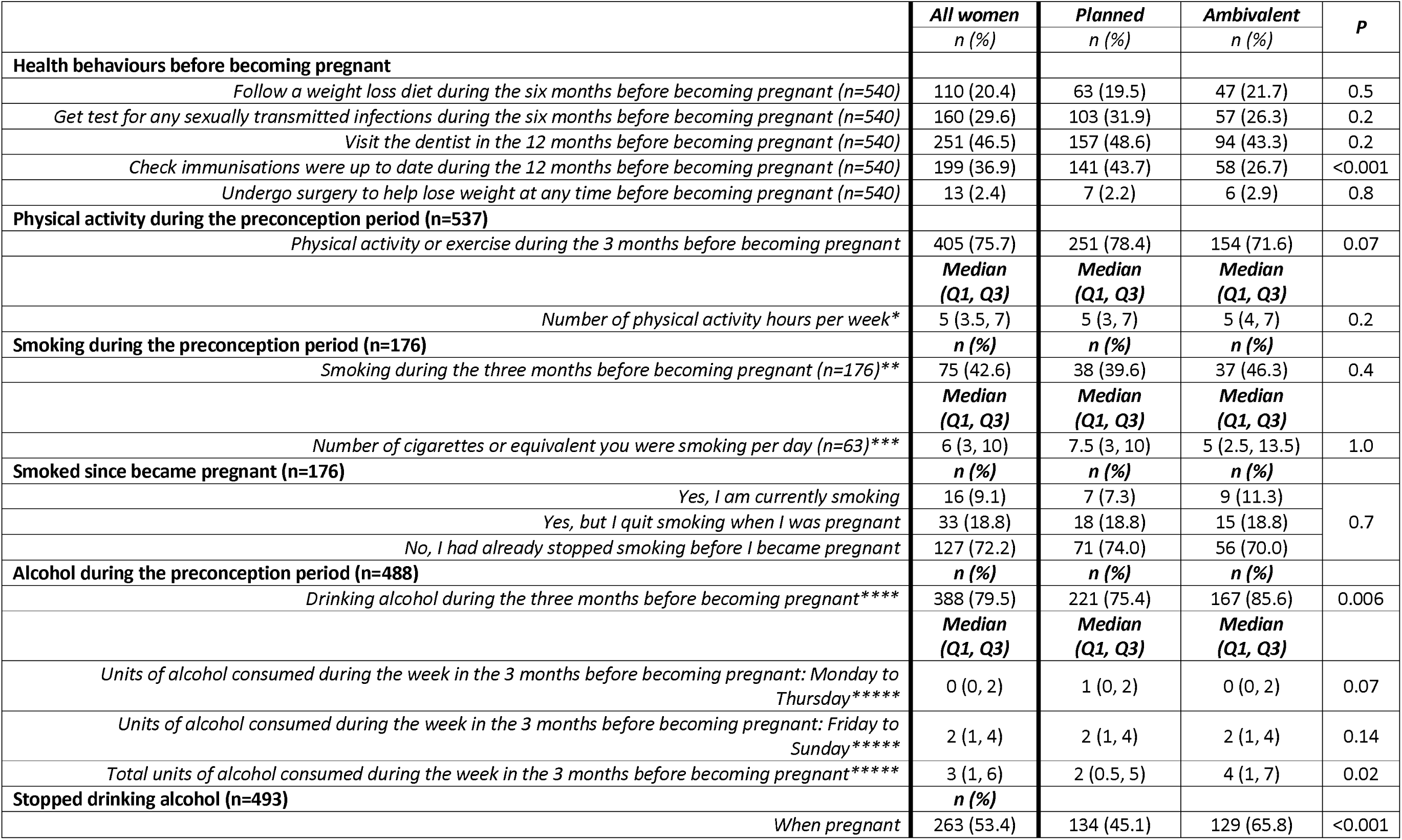
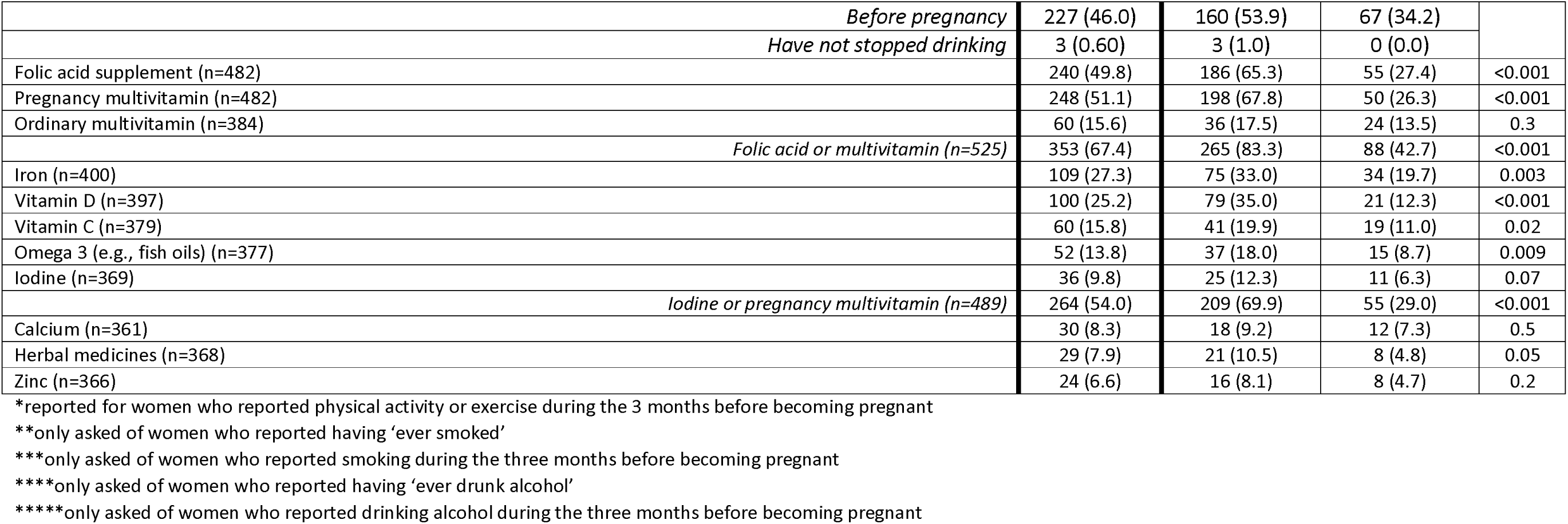
Preconception health behaviours.

**Table 5:** Association between BMI Classification and Physical activity or exercise, or following a weight loss diet before conception.

| BMI Classification | Physical activity or exercise in 3 months before conception (n=530) |  |  | Follow a weight loss diet in 6 months before conception (n=535) |  |  |
| --- | --- | --- | --- | --- | --- | --- |
|  | Yes | No | *p (V)* | Yes | No | *p (V)* |
|  | n (%) | n (%) |  | n (%) | n (%) |  |
| Underweight (n=14) | 8 (57.1) | 6 (42.9) | 0.001 (0.194) | 0 (0.0) | 15 (100.0) | <0.001 (0.220) |
| Normal weight (n=250) | 202 (80.8) | 48 (19.2) |  | 36 (14.3) | 215 (85.7) |  |
| Pre-obesity (n=141) | 107 (75.9) | 34 (24.1) |  | 33 (23.1) | 110 (76.9) |  |
| Obesity Class I (n=68) | 47 (69.1) | 21 (30.9) |  | 27 (39.1) | 42 (60.9) |  |
| Obesity Class II (n=31) | 24 (77.4) | 7 (22.6) |  | 8 (25.8) | 23 (74.2) |  |
| Obesity Class III (n=26) | 12 (46.2) | 14 (53.9) |  | 7 (26.9) | 19 (73.1) |  |

Smoking during the three months prior to pregnancy was reported by 42.6% of participants who reported having ‘ever smoked’, while drinking alcohol was reported over the same period by 79.5% of women who reported having ‘ever drunk alcohol’. The majority (72.2%) of participants who had smoked in the three months prior to conception had stopped smoking before becoming pregnant. Participants who were classified as having a ‘planned’ pregnancy consumed two units of alcohol per week while those with ‘ambivalent/unplanned’ pregnancy reported four units per week (p=0.02). Almost all participants who reported drinking before pregnancy had stopped at the time of survey completion, with 46.0% stopping before conception and 53.4% when they were pregnant. Women who reported drinking alcohol at any time during pregnancy consumed a median of five units of alcohol per week (Q1: 2, Q3, 8), with 25% consuming eight units or more per week, or two or more units per weekday and 75% consuming three or more units on a weekend day (data not shown).

Approximately half of all participants reported using a folic acid supplement (49.8%) or pregnancy multivitamin (51.1%) prior to pregnancy. When combined with those who used an ordinary multivitamin, 67.4% of participants used a supplement that likely contained folic acid in the preconception period. Analysis found higher prevalence of use of a folic acid-containing supplement prior to pregnancy among participants who ‘planned’ (83.3%) compared to those who were ‘ambivalent/unplanned’ (42.7%) about their pregnancy (p<0.001). More than half of participants (54.0%) used either an iodine supplement or a pregnancy multivitamin prior to becoming pregnant. When compared to women with an ‘ambivalent/unplanned’ pregnancy, a greater proportion of women with a ‘planned’ pregnancy reported intake of folic acid, iron, vitamin D, vitamin C, omega 3, zinc and pregnancy multivitamins (p≤0.05). For those participants using specific supplements, the duration of use prior to pregnancy varied with most users reporting taking a supplement for three to six months or for more than six months prior to becoming pregnant (Table 6). Among women who were taking a supplement, no statistically significant relationship between duration of use of that supplement and degree of pregnancy planning was found.

**TABLE 6:** DURATION OF SUPPLEMENT USE PRIOR TO CURRENT PREGNANCY, BY CATEGORY OF PREGNANCY PLANNING.

|  | All supplement users |  |  |  | Planned |  |  |  | Ambivalent/ Unplanned |  |  |  | *p* |
| --- | --- | --- | --- | --- | --- | --- | --- | --- | --- | --- | --- | --- | --- |
|  | <1<br>month | 1-3<br>months | 3-6<br>months | >6<br>months | <1<br>month | 1-3<br>months | 3-6<br>months | >6<br>months | <1<br>month | 1-3<br>months | 3-6<br>months | >6<br>months |  |
| Folic acid supplement (n=179) | 14<br>(7.8) | 60<br>(33.5) | 67<br>(37.4) | 38<br>(21.2) | 11<br>(7.6) | 47<br>(32.6) | 54<br>(37.5) | 32<br>(22.2) | 3 (8.6) | 13<br>(37.1) | 13<br>(37.1) | 6<br>(17.1) | 0.9 |
| Pregnancy multivitamin (n=209) | 18<br>(8.6) | 66<br>(31.6) | 75<br>(35.9) | 50<br>(23.9) | 14<br>(18.1) | 53<br>(30.5) | 66<br>(37.9) | 41<br>(23.6) | 4<br>(11.4) | 13<br>(37.1) | 9<br>(25.7) | 9<br>(25.7) | 0.6 |
| Ordinary multivitamin (n=27) | 1 (3.7) | 2 (7.4) | 7<br>(25.9) | 17<br>(62.9) | 1 (6.7) | 1 (6.7) | 5<br>(33.3) | 8<br>(53.3) | 0 (0.0) | 1 (8.3) | 2<br>(16.7) | 9<br>(75.0) | 0.6 |
| *Folic acid or multivitamin (n=296)* | 22<br>(7.4) | 89<br>(30.1) | 109<br>(36.8) | 76<br>(25.7) | 16<br>(6.9) | 70<br>(30.0) | 89<br>(38.2) | 58<br>(24.9) | 6 (9.5) | 19<br>(30.2) | 20<br>(31.8) | 18<br>(28.6) | 0.7 |
| Iron (n=74) | 2 (2.7) | 15<br>(20.3) | 22<br>(29.7) | 35<br>(47.3) | 1 (1.8) | 14<br>(24.6) | 19<br>(33.3) | 23<br>(40.4) | 1 (5.9) | 1 (5.9) | 3<br>(17.7) | 12<br>(70.6) | 0.08 |
| Vitamin D (n=73) | 7 (9.6) | 18<br>(24.7) | 23<br>(31.5) | 25<br>(34.3) | 4 (6.7) | 16<br>(26.7) | 19<br>(31.7) | 21<br>(35.0) | 3<br>(23.1) | 2<br>(15.4) | 4<br>(30.8) | 4<br>(30.8) | 0.3 |
| Omega 3 (e.g., fish oils) (n=37) | 2 (5.4) | 7<br>(18.9) | 14<br>(37.8) | 14<br>(37.8) | 1 (3.7) | 7<br>(25.9) | 10<br>(37.0) | 9<br>(33.3) | 1<br>(10.0) | 0 (0.0) | 4<br>(40.0) | 5<br>(50.0) | 0.3 |
| Iodine (n=24) | 1 (4.2) | 7<br>(29.2) | 9<br>(37.5) | 7<br>(29.2) | 0 (0.0) | 4<br>(23.5) | 7<br>(41.2) | 6<br>(35.3) | 1<br>(14.3) | 3<br>(42.9) | 2<br>(28.6) | 1<br>(14.3) | 0.4 |
| *Iodine or pregnancy multivitamin (n=223)* | 18<br>(8.1) | 68<br>(30.5) | 82<br>(36.8) | 55<br>(24.7) | 14<br>(7.6) | 54<br>(29.4) | 71<br>(38.6) | 45<br>(24.5) | 4<br>(10.3) | 14<br>(35.9) | 11<br>(28.2) | 10<br>(25.6) | 0.6 |

### Characteristics of women with planned pregnancies

The regression analysis retained variables describing the characteristics of women with planned pregnancies compared to women with ‘ambivalent/unplanned’ pregnancies (Table 7). Women who ‘planned’ a pregnancy were less likely to have poor or fair health rather than excellent health (AOR 0.3, p=0.02) compared to women with ‘ambivalent/unplanned’ pregnancy. ‘Planned’ pregnant women were more likely to be classified as ‘pre-obesity’ rather than ‘normal weight’ (adjusted odds ratio (AOR) 2.3, p=0.03) and to report not using a contraceptive within six months before pregnancy, compared to the other women (AOR 4.6, p<0.001). Women with ‘planned’ pregnancy were also less likely than women with ‘ambivalent/unplanned’ pregnancy to drink alcohol during the three months before becoming pregnant (AOR 0.4, p=0.04) and more likely to use a folic acid (AOR 3.0, p=0.002) or vitamin D supplement (AOR 2.4, p=0.03). Figure 1 presents the mapping of the participants’ preconception behaviours investigated in this study to the RACGP preconception care guidelines including where risk factors differ between women with ‘planned’ and ‘ambivalent/unplanned’ pregnancies.

**Figure 1:**
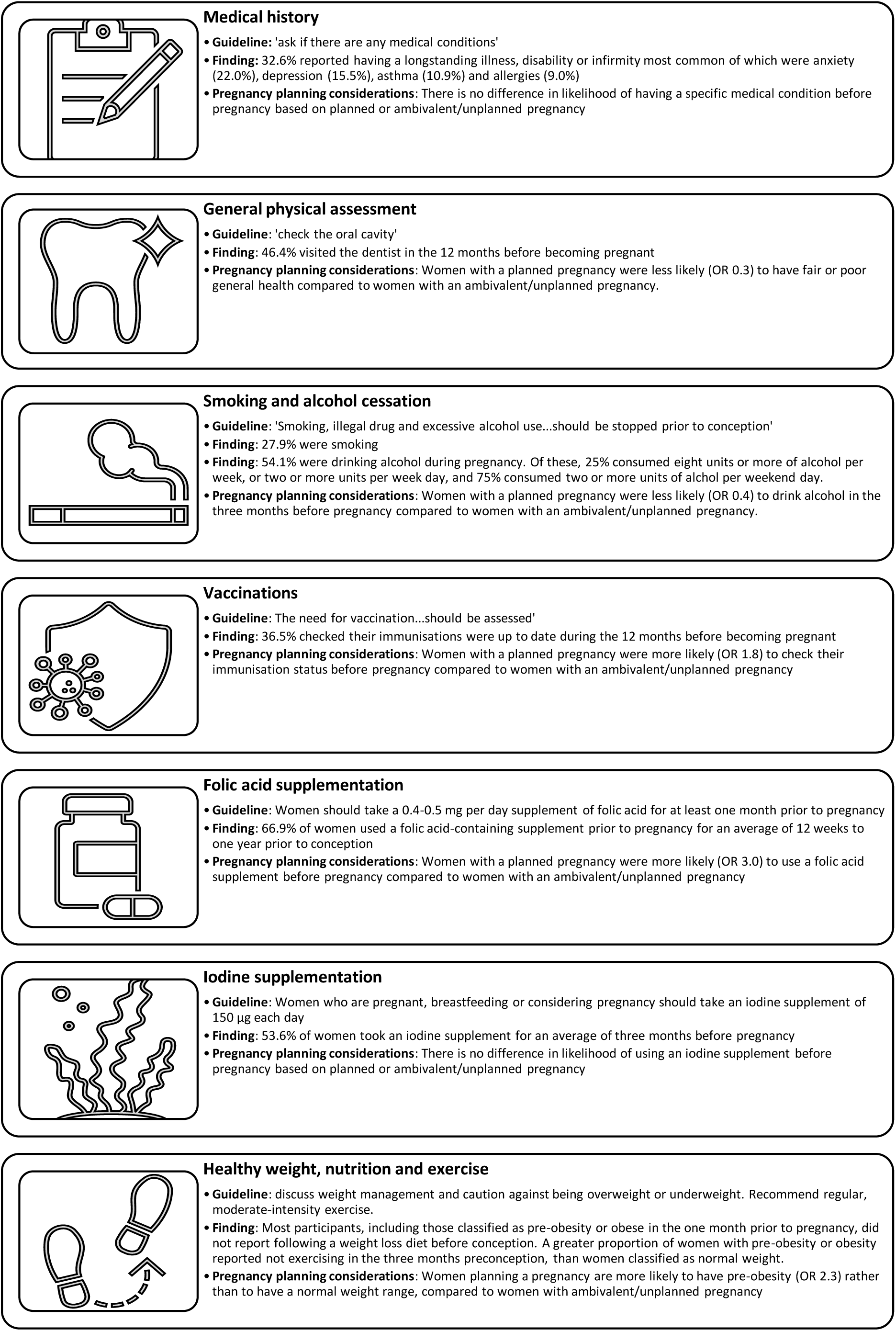
Women’s preconception care behaviours compared with the Royal Australian College of General Practitioner Guidelines [13] for Preventive Activities Prior to Pregnancy, including considerations based on Women’s pregnancy Planning

**Table 7:** Characteristics likely to be associated with reporting planned pregnancy, compared with ambivalent/unplanned pregnancy, based on multivariable regression analysis (n = 281)

| Characteristic category | Characteristics* | aOR (95% CI) | p |
| --- | --- | --- | --- |
| Demographics | Financial |  |  |
|  | *It is easy* | Ref | - |
|  | *It is not too bad* | 0.8 (0.4-1.6) | 0.5 |
|  | *It is difficult some of the time* | 0.5 (0.2-1.1) | 0.08 |
|  | *It is impossible/difficult all the time* | 0.5 (0.1-2.1) | 0.4 |
| Health history | General health status |  |  |
|  | *Excellent* | Ref | - |
|  | *Good* | 0.6 (0.3-1.5) | 0.3 |
|  | *Fair/Poor* | 0.3 (0.1-0.8) | 0.02 |
|  | BMI classification |  |  |
|  | *Normal weight* | Ref | - |
|  | *Underweight* | 2.2 (0.4-13.6) | 0.4 |
|  | *Pre-obesity* | 2.3 (1.1-4.8) | 0.03 |
|  | *Obesity class I* | 0.8 (0.3-2.0) | 0.6 |
|  | *Obesity class II* | 1.5 (0.4-5.2) | 0.5 |
|  | *Obesity class III* | 1.3 (0.3-4.9) | 1.0 |
|  | Contraception use 6 months before pregnancy |  |  |
|  | *Yes* | Ref | - |
|  | *No – I was trying to conceive* | 4.6 (2.1-10.1) | <0.001 |
|  | *No – I was not trying to conceive* | 0.7 (0.3-1.4) | 0.3 |
| Preconception health behaviour | Check immunisations were up to date during the 12 months before becoming pregnant | 1.8 (0.97-3.5) | 0.06 |
|  | Drinking alcohol during the three months before becoming pregnant | 0.4 (0.2-0.9) | 0.04 |
|  | Use of folic acid supplement | 3.0 (1.5-6.1) | 0.002 |
|  | Use of pregnancy multivitamin | 2.0 (0.97-4.0) | 0.06 |
|  | Use of Vitamin D | 2.4 (1.1-5.4) | 0.03 |

## DISCUSSION

Our national survey identifies 59% of pregnancies in Australia in 2020-21 were planned, which is lower than previous estimates. For example, 67.6% of women presenting to antenatal care in Sydney in 2010-11 had planned pregnancies [22], while 74% of pregnancies managed through maternity services in Victoria in 2017 were planned [12]. More recently, Black et al [23], found that 70.6% of women receiving antenatal care in two hospitals in Sydney in 2019-2020 could be categorised through the LMUP as having planned pregnancies. There are several reasons why our analysis may show a lower rate of ‘planned’ pregnancy. Firstly, we collected data during the COVID pandemic, which affected people’s childbearing decisions and access to contraception and abortion services [31-33]. Furthermore, abortion law reform was underway immediately prior to and during the time captured through the survey responses in five Australian states and territories (Queensland, New South Wales, South Australia, Western Australia, and Northern Territory [34]) and these changes may have meant that women with unplanned pregnancies that did not wish to continue their pregnancy may have been able to access abortion services more readily than in previous years. Such external factors may have influenced the proportion of planned vs unplanned pregnancies in our study and requires further research and monitoring. Secondly, previous studies recruited from antenatal care, whereas we recruited from social media giving an opportunity to include women who were not planning to continue their pregnancy. However the low levels of unplanned pregnancy (2.1%) in our sample are similar to that seen in the antenatal studies [25, 35], suggesting that we may have missed some women with unplanned pregnancies—had all pregnant women been recruited this figure may be as high as 9% [36]. This difference may be due to women being unlikely to enrol in a study about pregnancy if they are planning to terminate. Finally, the error in the online survey item design for the LMUP resulted in participants only being able to select one health behaviour rather than all relevant behaviours from the list provided. This may have led to a slight underestimate of the levels of pregnancy planning.

We have shown that pregnancy intention is associated with a range of socio-demographic characteristics, including age, parity, relationship status, education level and financial status, which is in keeping with the wider literature on the determinants of pregnancy intention [12, 36-39]. In general, women with the ‘planned’ pregnancies in our study tended to be aged in their 30s, or married, or employed with a university qualification or have good financial circumstances. This resonates strongly with the factors commonly reported as influencing individuals’ decisions around if or when to have a child [40]. Our study also found that ‘planned’ pregnancy was associated with earlier pregnancy recognition, which is important as such recognition often facilitates earlier uptake of antenatal care and can improve outcomes [41, 42]. Previous health and health behaviours - such as smoking, alcohol consumption and vitamin use - vary according to the level of pregnancy intention, with lower levels of risk behaviours (e.g., alcohol or tobacco use) or pre-existing mental health conditions occurring in women with ‘planned’ pregnancies. This trend is also in keeping with previous literature [12, 36, 43], though our findings of better self-reported general health are novel. This new insight may require an increasingly salutogenic approach to future preconception health messaging to inspire ‘well’ populations to pay attention to address problematic preconception health behaviours. Such a need for salutogenic preconception health messaging is further reinforced by findings from a UK study which found more than half of a sample of women who were currently pregnant or preparing for pregnancy reported not engaging in preconception health behaviour change as they viewed themselves as ‘already healthy’ [44]. However, our study may also suggest that women who are not in good health may be less likely to consider the time is right for a pregnancy. The lack of relationships between pregnancy planning and other categories of health conditions is likely due to the small number of participants identifying as having some of the health conditions (e.g. cardiometabolic and endocrine conditions) and consequent lack of statistical power. The multivariable model, built using a conceptual epidemiological hierarchy, confirmed that financial status, BMI, general health, and behaviours before pregnancy are all associated with pregnancy planning. Unusually variables such as age, relationship status and parity did not pass the likelihood ratio test and were able to be removed from the model after the health history variables were added. Once preconception health behaviours were included in the model, the level of education variable was also able to be removed. Ultimately, these outcomes of the regression analysis suggest that financial status is the most important distal determinant of pregnancy planning.

Our study shows that actions to improve health before pregnancy were relatively uncommon, though more likely in women who planned their pregnancies. The most common action was taking folic acid or a pregnancy multivitamin, which was even greater in planned pregnancies. It is encouraging to see relatively high levels of supplement use, suggesting that there is awareness of the importance of folate in the preconception period; a finding also reflected in a UK survey of women who were pregnant or planning pregnancy [44]. In contrast, routine maternity services data in England has found less than one quarter of pregnant women had taken a folic acid supplement before pregnancy. The fact that, despite decades of evidence and widespread recommendations, nearly 20% of planned pregnancies in our study were not protected by folate supplementation – and between 75% and 50% of women in UK research [34, 44] - remains disappointing. This finding should also be considered in light of emerging evidence regarding women potentially exceeding required levels of folate intake in preconception and pregnancy by taking supplements in countries, like Australia, with folic acid fortification of food [45]. Unfortunately, the reason why participants in our study did not use a folic acid supplement was not captured by our survey instrument and this issue requires further research. Iodine is also recommended before pregnancy in Australia [46] but only 9.8% of women reported taking iodine in preconception, potentially suggesting low levels of awareness. While this rate was higher among women who were planning a pregnancy, there was no significant relationship identified after accounting for confounders via logistic regression. Beyond supplement use, previous systematic mapping of the literature [47] highlights that studies investigating associations between pregnancy intention and preconception behaviours/psychological wellbeing gave insufficient attention to diet, exercise, and psychological factors (compared to supplementation) and rarely used a validated measure of pregnancy intention. Our study uses the validated Australian LMUP and has considered a range of preconception behaviours and this is a real strength. This analysis also found a weak to moderate association between BMI classification and engaging in preconception physical activity or following a preconception weight loss diet, but these relationships were not linear, and the regression analysis only found participants in the pre-obesity category were more likely to report planning a pregnancy. Given the levels of obesity in people of reproductive age in Australia [48], and the intergenerational transmission of obesity [6], factors linked with obesity such as diet, exercise and mental health require focused attention. Women with pre-obesity and obesity may require additional support to implement such health behaviours if they are individually appropriate, ideally whilst addressing the issue of weight stigma to which these groups are particularly vulnerable [49].

Such low levels of preconception preparations are not uncommon; contemporary data from Australia shows that less than half of women attending antenatal care had taken any steps to improve their health before pregnancy [23] and data from the UK shows that barely one in three took folic acid before pregnancy in 2018/19 [50]. Overall, our study findings highlighted the gap between the RACGP guidelines for preconception care [18] and the preconception health behaviours for women; further emphasising the potential critical role of health professionals and public health promotion agencies in increasing awareness in the full range of behaviour change requiring attention in the preconception period. These gaps may also be used to further inform current efforts towards priority setting for preconception health policy, research and practice in Australia [51]. Data such as those captured in this survey are not routinely available at a national level but, when available, such data can be incredibly useful to identify areas requiring additional public health attention as well as measuring progress where population-level health interventions are implemented. The UK has recently published its first preconception report card [50, 52] and Australia is making steps to do the same [51, 53]. Certainly, our study adds weight to existing calls for strengthened primary care that is both comprehensive and family-oriented, to improve maternal health before pregnancy to achieve better birth outcomes [54]. General practitioners, in particular, are primary care practitioners consulted by more than 80% of the population [55], and as the most common health professional seen by the public they are a well-positioned and largely untapped resource to deliver preconception care [56]. Overall, Australia urgently needs a primary care strategy for the delivery of preconception care that enables implementation of the RACGP recommendations and leverages the skills, capacity and reach of the full breadth of the primary care workforce to optimise continuity of care in the primary care setting [57, 58].

While it is important to highlight specific actions that individuals can undertake with regards to preconception health, it is also vital that there is also a focus upon raising a wider awareness of the opportunities, abilities and benefits of planning and preparing for pregnancy. There is also a need for structural barriers—such as out-of-pocket costs for health services and products, health literacy, and clear identification of health professionals trained to deliver preconception care—and the impact of the wider determinants of health to be addressed. This why a dual strategy towards preconception health has been proposed [52] and a community-based universal model of care that includes schools, digital health, social media, and other routes, while centring user preferences has been described as a route to this [59]. Such an approach will also help address the phenomenon seen in our data, where women describe their health as good overall, and yet often enter into pregnancy with a number of risk factors, possibly because they are unaware of the relationships between pregnancy planning, preconception health and fertility and pregnancy outcomes [60, 61].

### Limitations

The findings from our study should be interpreted within the context of the study’s limitations. Firstly, our participants were recruited through social media. This method provided a broader exposure to the target population than those allowed through previous study designs which recruited through health services. However, social media recruitment still has the potential to create a sampling bias by recruiting individuals with higher levels of education and other sociodemographic differences [62]. Future research may need to explicitly employ recruitment methods targeting populations with lower education levels to supplement the findings arising from this study. Another potential effect of sampling bias is the difference in age and state of residence between the study population and the Australian population. The study sample demonstrated a higher proportion of women aged 20 to 29 years compared to the national data, and it is possible that this could have resulted in the study findings deviating from true prevalence of pregnancy planning and preconception behaviours. However, as our multivariable analysis identified, age was not related to pregnancy planning this limitation may be minimal. The univariate associations identified from our analyses must be interpreted with caution, given their cross-sectional nature and the high likelihood of confounding, and as such we have primarily based our findings on the outcomes of the regression analysis. The error in the survey instrument item associated with the LMUP resulted in an incomplete LMUP score, but as the analysis was based on binary groups the impact of this error is likely to be minimal. It is also worth noting that some survey items capture general behaviour before pregnancy (e.g., see a dentist) and these behaviours may not necessarily represent an action taken specifically to prepare for pregnancy. This lack of differentiation between general health behaviours and intended preconception health behaviours may explain the lack of significant differences between such variables. Lastly, while we have reduced post hoc rationalization by asking participants about their preconception behaviours during pregnancy rather than after birth, there is a chance of some recall bias in that women were asked to remember behaviours from up to 24 months before they completed the survey. Overall, these limitations should be carefully considered when interpreting the results and drawing conclusions to inform future research, practice and policy.

## CONCLUSIONS

While the majority of women may report some degree of pregnancy planning, this national study suggests a lower proportion of pregnancy planning when sampling pregnant women in the community compared to women accessing antenatal services. Even among women who have planned their pregnancy, the limited actions they have taken to improve their health prior to pregnancy is concerning. The most common health behaviour was use of a vitamin supplement such as folate, but there are a wider range of important health behaviour changes that may be needed for women to achieve optimal preconception health. With this in mind, there is a pressing need to develop more comprehensive preconception health interventions for primary care and health promotion to support women to enact positive behaviour change prior to pregnancy. In addition to promoting specific actions that individuals can undertake with regards to preconception health, it is vital that the impact of structural barriers and wider determinants of health be adequately considered and addressed in terms of preconception health promotion, planning and policy. Such a broad approach can help strengthen attempts to improve pregnancy planning and preconception health.

## Data Availability

The datasets analysed during the current study are available from the corresponding author on reasonable request.

## ABBREVIATIONS

PCH: Preconception health
PCC: Preconception care
DOHaD: Developmental origins of health and disease
LMUP: London Measure of Unplanned Pregnancy
BMI: Body mass index
ANOVA: Analysis of variance
AOR: Adjusted odds ratio
RACGP: Royal Australian College of General Practitioners

## DECLARATIONS

### ETHICS APPROVAL AND CONSENT TO PARTICIPATE

This study was conducted in accordance with the principles of the Declaration of Helsinki. Informed consent was obtained from all participants prior to accessing survey questions. The study was reviewed and approved by the University of Technology Sydney Medical and Health Research Ethics Committee (#ETHI21-6401).

### CONSENT FOR PUBLICATION

Not applicable.

### AUTHORS’ CONTRIBUTIONS

AS conceived the research questions and designed the study. AS, DS, EM, JH and WB designed the study instrument. AS, DS and JH developed the analysis plan, and drafted the methods, and results. AS, JH, AL, EM, JA, WB and DS contributed to drafting all other manuscript sections and reviewed and approved the final draft.

## ACKNOWLEDGEMENTS

We would like to acknowledge Associate Professor Kris Rogers (School of Public Health, University of Technology Sydney) for his expert statistical advice and guidance for this study.

