## Supplementary File 1 for "WOMEN’S PRECONCEPTION HEALTH, PLANNING, AND BEHAVIOURS: A CROSS-SECTIONAL SURVEY OF PREGNANT WOMEN IN AUSTRALIA"

Women's Pre-pregnancy Health Information and Health Behaviours

#### Default Question Block

**Welcome to the Survey of Women's Pre-pregnancy Health Information and Behaviours**

Have you received and read the Participant Information Sheet for this study

Yes

No

***Preconception care beliefs, attitudes and behaviours of women and men***
***(HREC# ETH20-4726)***

WHO IS DOING THE RESEARCH?
My name is Dr Amie Steel and I am an academic at the University of Technology Sydney (UTS).  My research team includes Dr Erica McIntyre (UTS), Dr Jane Frawley (UTS), Professor Jon Adams (UTS), Dr Danielle Schoenaker (University of Wollongong), Dr Anna Gavine (University of Dundee), Ms Adina Lang (Monash University) and Dr Jenny Hall (University College London).

WHAT IS THIS RESEARCH ABOUT?

This research is to find out about the preconception beliefs, attitudes and behaviours of pregnant women and their reproductive partners.

FUNDING
Funding for this project has been received from the Faculty of Health at UTS.

WHY HAVE I BEEN ASKED?
You have been invited to participate in this study because you are a pregnant woman or the reproductive partner of a pregnant woman.

IF I SAY YES, WHAT WILL IT INVOLVE?
If you decide to participate, I will invite you to complete a survey about your beliefs, attitudes and behaviours in the time before you were pregnant and during your current pregnancy. The survey will take approximately 30 minutes to complete and can be filled out on your own device. You will be asked to provide your email address so that an email containing a link to the survey can be sent to you. A second email containing a link to an survey for your partner to complete will also be sent to the same email address and you are invited to share this link with your partner. Both your survey responses and the responses provided by your partner, should they choose to participate, also, will be linked through a shared ID code. This ID code will be linked to your email address in a database separate from your survey responses. No other identifying information, such as yours or your partner's name, postal address or phone number, will be collected in any aspect of this project.
If you are unable to complete the survey in one sitting, you will be able to save your responses and return to the survey on the same device at a later time. Once started, you will have up to 14 days to complete the survey.
Upon completing the survey, you will be provided a link to enter a competition to win a $100 voucher to spend online at fishpond.com.au in acknowledgement of your time. The details you provide to enter the competition will not be in anyway linked to your survey response or used for any purpose other than selecting the winner of the draw. No third party will have access to these details. After the winner is notified and the voucher is delivered, your details will be deleted.

ARE THERE ANY RISKS/INCONVENIENCE?
Yes, there are some risks or inconvenience.  You will be asked some personal health questions including about your pregnancy history. Some of these questions may cause emotional distress to some people.

DO I HAVE TO SAY YES?
Participation in this study is voluntary. It is completely up to you whether or not you decide to take part.

WHAT WILL HAPPEN IF I SAY NO?
If you decide not to participate, it will not affect your relationship with the researchers or the University of Technology Sydney, or your clinical care at the location where you were approached about this study. If you wish to withdraw from the study once it has started, you can do so at any time before you return the survey to the research team at the clinic where you were first told about the study, or via post.
However, it is not possible to withdraw your data from the study results after the survey items have been completed as all surveys will be anonymous.

CONFIDENTIALITY
By completing the survey, you consent to the research team collecting and using information about you for the research project. All this information will be treated confidentially. All survey responses will be anonymous and will not be able to be identified to you.  Your information will only be used for the purpose of this research project. Your survey will be matched to your partners survey via an unidentifiable code. We plan to publish the results in peer-reviewed scientific journals.

WHAT IF I HAVE CONCERNS OR A COMPLAINT?
If you have concerns about the research that you think I can help you with, please feel free to contact me on 0418 786 186 or

A copy of this information sheet will be included in the email you were sent with the link to this survey.

**NOTE:** This study has been approved in line with the University of Technology Sydney Human Research Ethics Committee [UTS HREC] guidelines.  If you have any concerns or complaints about any aspect of the conduct of this research, please contact the

Ethics Secretariat on ph.: +61 2 9514 2478 or], and quote the UTS HREC reference number (ETH20-4276). Any matter raised will be treated confidentially, investigated and you will be informed of the outcome.

This questionnaire will ask questions about your health, your knowledge of pre-pregnancy health, and any experiences relating to pre-pregnancy care that you may have had prior to your current pregnancy. You can either complete this now during your clinic visit or at a later stage. It will take about 30 minutes to complete. Any information you provide will remain anonymous.
**Note**: Participation in this study is voluntary. You are not required to participate even if your partner has chosen to participate.

**By completing the survey, you agree to the following:**

I have read the participant information sheet, or someone has read it to me in a language that I understand. I understand the purposes, process and risks of the research described. I have had an opportunity to ask questions and I am satisfied with the answers I have received. I have not completed this survey previously. I freely agree to participate in the research survey described and understand that I am free to withdraw at any time without affecting the health care I receive.

Yes

No

Please read each question carefully. Please answer every question to the best of your knowledge. If you are unsure about how to answer a question, mark the option that most closely describes your response.

Please note that you can take a break from responding to the survey and return at a later stage but you will need to access it with the same browser and device as you are currently using and complete the survey within 14 days.

#### Section 1: About You

##### What is your age (in years)?

##### What is your gender?

Female

Male

Transgender

Other

##### What is your residential postcode?

#### What is your relationship status?

Married

De facto

Not currently in a relationship

In a relationship but not living with my partner

Other (please give details

#### What is the highest qualification you have completed?

No formal qualification

Year 10 or equivalent

Year 12 or equivalent

Trade or apprenticeship

Certificate or diploma

University degree (e.g. Bachelor)

Higher university degree (e.g. Masters, PhD)

#### What best describes your employment status?

Full time work (35 hours or more per week)

Part time work (less than 35 hours per week)

Casual or temporary work (irregular hours)

Currently looking for work

Not currently in the paid workforce, nor looking

#### How do you manage financially at the moment?

It is impossible

It is difficult all the time

It is difficult some of the time

It is not too bad

It is easy

#### Do you currently have private health insurance?

No

Yes, but no obstetrics cover

Yes, with obstetrics cover

#### Do you currently have a Health Care Card?

Yes

No

#### How tall are you?

#### What was your weight one month before pregnancy? (in kilograms)

#### What is your current weight? (in kilograms)

#### What other adults do you live with?

husband

husband and (your/your husband's) parents

partner

partner and (your/your partner's) parents

alone

your parents

other relatives or friends

other (e.g. living in accommodation provided with your job, etc)

#### Is English your first language?

Yes

No, please list your first language:

Do you identify as Aboriginal or Torres Strait Islander?

Yes

No

I would rather not say

Were you born in Australia?

Yes

No, please give details:

#### Block 2

The remainder of this survey relates to your current pregnancy.

### Section 2: Your current pregnancy

How many weeks pregnant are you now?

What is the estimated due date for your pregnancy?

How many weeks pregnant were you when you found out you were pregnant?

How did you find out you were pregnant?

Home pregnancy test

From a health professional

Other

#### Is this current pregnancy the result of you or your partner receiving fertility treatments?

Yes

No

#### How long before trying to become pregnancy were you thinking about having a baby?

I was not trying to become pregnant

Less than 1 month

Between 1 and 2 months

Between 3 and 5 months

Between 6 and 12 months

More than 12 months

#### Are there any other circumstances related to this pregnancy which you would like to share?

#### Below are some questions that ask about your circumstances and feelings around the time you became pregnant. Please think of your current pregnancy when answering the questions below (please tick the statement that most applies to you):

#### In the month that I became pregnant...

I/we were not using contraception (including no need for contraception, e.g. fertility treatment, female partner, etc.)

I/we were using contraception, but not on every occasion

I/we always used contraception, but knew that the method had failed (i.e. broke, moved, came off, came out, not worked etc) at least once

I/we always used contraception

#### In terms of becoming a mother (first time or again), I feel that my pregnancy happened at the...

Right time

Ok, but not quite right time

Wrong time

#### Just before I became pregnant...

I intended to get pregnant

My intentions kept changing

I did not intend to get pregnant

#### Just before I became pregnant...

I wanted to have a baby

I had mixed feelings about having a baby

I did not want to have a baby

#### In the next question, we use the word 'partner'—this might be (or have been) your husband/wife/civil partner, a partner you live with, a partner who lives elsewhere, someone you've had sex with once or twice, or a parenting (non-romantic partner)

#### Before I became pregnant...

My partner and I had agreed that we would like me to be pregnant

My partner and I had discussed having children together, but hadn't agreed for me to get pregnant

We never discussed having children together

I chose to become pregnant without a partner

### Section 3: Your Health Behaviours

#### Before you became pregnant, did you do anything to improve your health in preparation for your pregnancy?

Took folic acid

Took iodine

Took Vitamin D

Stopped or cut down smoking

Stopped or cut down drinking alcohol

Ate more healthily

Sought medical/health advice

Took some other action, please describe

I did not do any of the above before my pregnancy

Some women take vitamins or supplements BEFORE they are pregnant.

Did you take any vitamins or supplements BEFORE you became pregnant?
(a) If ‘No’, please tick the box
(b) If ‘Yes’, how many weeks before you became pregnant did you start taking each vitamin or supplement?
(c) If ‘Yes’, how often did you take each vitamin or supplement?

| | (a) | (b) | (c) | | | |
|---|---|---|---|---|---|---|
| | No | Number of weeks before pregnant | Every day | Most days | Occasionally | Other |
| Folic acid | | | | | | |
| Pregnancy multivitamin | | | | | | |
| Ordinary multivitamin | | | | | | |
| Vitamin D | | | | | | |
| Iron | | | | | | |
| Omega 3 (e.g. fish oils) | | | | | | |
| Vitamin C | | | | | | |
| Zinc | | | | | | |

| | (a) | (b) | (c) | | | |
|---|---|---|---|---|---|---|
| | No | Number of weeks before pregnant | Every day | Most days | Occasionally | Other |
| Iodine | ○ | | ○ | ○ | ○ | ○ |
| Calcium | ○ | | ○ | ○ | ○ | ○ |
| Herbal medicines | ○ | | ○ | ○ | ○ | ○ |
| Other (please give details) | ○ | | ○ | ○ | ○ | ○ |
| Other (please give details) | ○ | | ○ | ○ | ○ | ○ |

Some women take vitamins or supplements WHEN they are pregnant.

(a) Have you taken any vitamins or supplements while you have been pregnant? (Please tick the box for each vitamin or supplement that applies)
If yes..
(b) when did you start taking each vitamin or supplement? Before or during this pregnancy?
(c) How often do you/did you take each vitamin or supplement?
(d) Are you still taking them?
(e) If you are not still taking them, when did you stop?

| | (a) | (b) | (b) | (c) |
|---|---|---|---|---|

| | (a) | | (b) | (b) | (c) | | | | |
|---|---|---|---|---|---|---|---|---|---|
| | Yes | No | BEFORE I was pregnant | Number of weeks of pregnancy | Every day | Most days | Occasionally | Other | Y |
| Folic acid | | | | | | | | | |
| Pregnancy multivitamin | | | | | | | | | |
| Ordinary multivitamin | | | | | | | | | |
| Vitamin D | | | | | | | | | |
| Iron | | | | | | | | | |
| Omega 3 (e.g. fish oils) | | | | | | | | | |
| Vitamin C | | | | | | | | | |
| Zinc | | | | | | | | | |
| Iodine | | | | | | | | | |
| Calcium | | | | | | | | | |
| Herbal medicines | | | | | | | | | |
| Other (please give details) | | | | | | | | | |

Yes No BEFORE I was pregnant Number of weeks of pregnancy Every day Most days Occasionally Other Y

| | (a) | | (b) | (b) | (c) | | | | |
|---|---|---|---|---|---|---|---|---|---|
| | Yes | No | BEFORE I was pregnant | Number of weeks of pregnancy | Every day | Most days | Occasionally | Other | Y |
| Other (please give details) | ○ | ○ | ○ | | ○ | ○ | ○ | ○ | ( |

Some women take medications to manage health complaints before and during pregnancy.

(a) Please list any medications you have taken in the 3 months before you became pregnant, and if you have taken them since you became pregnant. Include any pharmaceutical or non-pharmaceutical medications (e.g. herbal medicines) not already listed by you in this questionnaire. Include tablets, creams, drops, injections and inhalers either prescribed for you or bought over the counter. You can put the type of medication if you do not know the name.

(b) Please identify any medications that have been reviewed or discussed with a health professional either before you became pregnant or since you became pregnant

| | (a) | (a) | | (b) | | | | |
|---|---|---|---|---|---|---|---|---|
| | Please write | Taken BEFORE being pregnant | Taken SINCE being pregnant | Reviewed BEFORE being pregnant | Reviewed SINCE being pregnant | GP | Midwife | N |

| | (a) | (a) | | (b) | | | | |
|---|---|---|---|---|---|---|---|---|
| | Please write | Taken BEFORE being pregnant | Taken SINCE being pregnant | Reviewed BEFORE being pregnant | Reviewed SINCE being pregnant | GP | Midwife | N |
| Medication 1 (please write) ☐ | ☐ | ☐ | ☐ | ☐ | ☐ | ☐ | ☐ | |
| Medication 2 (please write) ☐ | ☐ | ☐ | ☐ | ☐ | ☐ | ☐ | ☐ | |
| Medication 3 (please write) ☐ | ☐ | ☐ | ☐ | ☐ | ☐ | ☐ | ☐ | |
| Medication 4 (please write) ☐ | ☐ | ☐ | ☐ | ☐ | ☐ | ☐ | ☐ | |
| Medication 5 (please write) ☐ | ☐ | ☐ | ☐ | ☐ | ☐ | ☐ | ☐ | |

| | (a) | (a) | | (b) | | | | |
|---|---|---|---|---|---|---|---|---|
| | Please write | Taken BEFORE being pregnant | Taken SINCE being pregnant | Reviewed BEFORE being pregnant | Reviewed SINCE being pregnant | GP | Midwife | N |
| Medication 6 (please write) ☐ | ☐ | ☐ | ☐ | ☐ | ☐ | ☐ | ☐ | |
| Medication 7 (please write) ☐ | ☐ | ☐ | ☐ | ☐ | ☐ | ☐ | ☐ | |

In the <u>3 months before</u> you became pregnant were you seeing any health professionals for ongoing assistance with your health (e.g. doctor for medical condition, counsellor for depression etc). (*If none, please write 'none'*)

| | Reason for visit (please write) |
|---|---|
| Type of health professional (please write) ☐ | ☐ |

| | Reason for visit (please write) |
|---|---|
| Type of health professional (please write) | |

#### Did you use any contraception in the 6 months before you became pregnant?

Yes

No—I was trying to conceive

No—I was not trying to conceive

#### Did you use any of the following methods of contraception in the 6 months before you became pregnant (*Please select all that apply*)

| | Never | Some of the time | Every time |
|---|---|---|---|
| Withdrawal | ○ | ○ | ○ |
| Condoms | ○ | ○ | ○ |
| Cap/diaphragm | ○ | ○ | ○ |

| | Never | Some of the time | Every time |
|---|---|---|---|
| Contraceptive patch | ○ | ○ | ○ |
| Abstinence (i.e. not having sex) | ○ | ○ | ○ |
| Emergency contraception (e.g. Levonelle, emergency IUD) | ○ | ○ | ○ |
| Intercourse timed around ovulation (e.g. Billings method) | ○ | ○ | ○ |
| Other (please give details) [ ] | ○ | ○ | ○ |

#### Did you use any of the following methods of contraception in the 3 months before you became pregnant (*Please tick all that apply*)

Contraceptive injection (e.g Depo-provera)

Contraceptive implant

Intrauterine device (IUD or coil)

Intrauterine system (IUS e.g. Mirena)

Birth control pills (oral contraceptives)

#### If you stopped using contraception to try and get pregnant, how long (in *weeks* or *months*) was it before you became pregnant?

Weeks

OR Months

#### Did you have an IUD or implant removed to try and get pregnant?

Yes

No

#### When the IUD or implant was removed, were you given any pre-pregnancy health and care information?

Yes, for me only

Yes, for me and my partner

No

### Did you do any of the following before you became pregnant?

Follow a weight loss diet during the 6 months before you became pregnant

Get tested for any sexually transmitted infections (e.g. chlamydia) during the 6 months before you became pregnant

Visit the dentist during the 12 months before you became pregnant

Check that your immunisations were up to date during the 12 months before you became pregnant

Undergo surgery to help you lose weight (e.g. bariatric surgery, gastric band, gastric balloon, gastric bypass) at any time before you became pregnant

### Were you doing any physical activity or exercise during the 3 months before you became pregnant?

Yes - please provide details in the next question

No

### Physical activity details...

Number of hours per week

Type(s) of exercise/activity

#### Have you done any physical activity or exercise <u>since</u> you became pregnant?

Yes, currently - please provide details below

Yes, but I stopped when I was pregnant (please provide week of pregnancy)

No

#### Physical activity details...

Number of hours per week

Type(s) of exercise/activity

#### Have you ever smoked?

Yes

No

#### Which of the following have you smoked?

Cigarettes

E-cigarettes

Other (please give details)

#### Did you smoke in the <u>3 months before</u> you became pregnant?

No

Yes, - please write the number of cigarettes or equivalent you were smoking per day

#### Have you smoked <u>since</u> you became pregnant?

Yes, I am smoking (please write number of cigarettes per day)

Yes, but I quit smoking when I was pregnant (please write week of pregnancy you quit)

No, I had already stopped smoking before I became pregnant

#### Did anyone (e.g. partner, family, friends, work colleagues) smoke indoors where you lived or worked before you became pregnant?

No

Yes (please give details)

#### Has anyone (e.g. partner, family, friends, work colleagues) smoked indoors where you lived or worked since you became pregnant?

No

Yes (please give details)

#### Have you ever drunk alcohol?

Yes

No

On average, how many units of alcohol did you have during the week in the 3 months before you became pregnant? How many units of alcohol did you have on the weekend? (*if none please state '0'*).

Note: a unit of alcohol is equivalent to half a pint of beer, 1 small glass of wine, or a single measure of spirits)

Units of alcohol during the week (Monday to Thursday)

Units of alcohol during the weekend (Friday to Sunday)

When did you stop drinking alcohol?

I stopped drinking when I was pregnant (please write the week of pregnancy)

I stopped before pregnancy

I have not stopped drinking alcohol

On average, how many units of alcohol are you currently drinking during the week? How many units of alcohol do you drink on the weekend? (*if none please state '0'*).

Note: a unit of alcohol is equivalent to half a pint of beer, 1 small glass of wine, or a single measure of spirits)

Units of alcohol during the week (Monday to Thursday)

Units of alcohol during the weekend (Friday to Sunday)

Think back on the whole of your pregnancy, including the first weeks before you knew you became pregnant. How many times did you consume five drinks or more in a single occasion (half a pint of beer, 1 small glass of wine, or a single measure of spirits) over this period? (*if none please put '0'*)

Number of times:

I don't remember/I don't know

Please answer each of the following questions by moving the marker to the number that best describes your opinion. Some of the questions may appear to be similar, but they do address somewhat different issues.

| Definitely true | | | | | Definitely false | | |
|---|---|---|---|---|---|---|---|
| 0 | 1 | 2 | 3 | 4 | 5 | 6 | 7 |

| | Definitely true | | | | | | Definitely false |
|---|---|---|---|---|---|---|---|
| | 0 | 1 | 2 | 3 | 4 | 5 | 6 | 7 |
| I exercised regularly for at least 6 months before becoming pregnant | | | | | | | | |
| I ate a healthy diet for at least 6 months before becoming pregnant | | | | | | | | |
| I avoided alcohol for at least 6 months before becoming pregnant | | | | | | | | |

Please answer each of the following questions by moving the marker to the number that best describes your opinion. Some of the questions may appear to be similar, but they do address somewhat different issues.

| | Important | | | | | | Not important |
|---|---|---|---|---|---|---|---|
| | 0 | 1 | 2 | 3 | 4 | 5 | 6 | 7 |
| My doing regular exercise during the 6 months prior to becoming pregnant is: | | | | | | | | |
| My eating a healthy diet during the 6 months prior to becoming pregnant is: | | | | | | | | |
| My avoiding alcohol during the 6 months prior to becoming pregnant is: | | | | | | | | |

Please answer each of the following questions by moving the marker to the number that best describes your opinion. Some of the questions may appear to be similar, but they do address somewhat different issues.

| | Pleasant | | | | | | Unpleasant | |
|---|---|---|---|---|---|---|---|---|
| | 0 | 1 | 2 | 3 | 4 | 5 | 6 | 7 |
| My doing regular exercise during the 6 months prior to becoming pregnant is: | | | | | | | | |
| My eating a healthy diet during the 6 months prior to becoming pregnant is: | | | | | | | | |
| My avoiding alcohol during the 6 months prior to becoming pregnant is: | | | | | | | | |

Please answer each of the following questions by moving the marker to the number that best describes your opinion. Some of the questions may appear to be similar, but they do address somewhat different issues.

| | Good | | | | | | | Bad |
|---|---|---|---|---|---|---|---|---|
| | 0 | 1 | 2 | 3 | 4 | 5 | 6 | 7 |
| My doing regular exercise during the 6 months prior to becoming pregnant is: | | | | | | | | |

| | Good | | | | | | Bad |
|---|---|---|---|---|---|---|---|
| | 0 | 1 | 2 | 3 | 4 | 5 | 6 | 7 |
| My eating a healthy diet during the 6 months prior to becoming pregnant is: | | | | | | | | |
| My avoiding alcohol during the 6 months prior to becoming pregnant is: | | | | | | | | |

Please answer each of the following questions by moving the marker to the number that best describes your opinion. Some of the questions may appear to be similar, but they do address somewhat different issues.

| | Extremely likely | | | | | | Extremely unlikely | |
|---|---|---|---|---|---|---|---|---|
| | 0 | 1 | 2 | 3 | 4 | 5 | 6 | 7 |
| Most people like me exercise regularly for 6 months before becoming pregnant | | | | | | | | |
| Most people like me eat a healthy diet for 6 months before becoming pregnant | | | | | | | | |
| Most people like me avoid alcohol for 6 months before becoming pregnant | | | | | | | | |

| | Extremely likely | | | | | Extremely unlikely | | | |
|---|---|---|---|---|---|---|---|---|---|
| | 0 | 1 | 2 | 3 | 4 | 5 | 6 | 7 | |
| If I maintain a healthy weight for 6 months before becoming pregnant my child will be healthy | | | | | | | | | ☐ |
| If I avoid alcohol for 6 months before becoming pregnant my child will be healthy | | | | | | | | | ☐ |
| If I exercise regularly for 6 months before becoming pregnant my child will be healthy | | | | | | | | | ☐ |
| If I eat a healthy diet for 6 months before becoming pregnant my child will be healthy | | | | | | | | | ☐ |
| If I increase folic acid intake for 6 months before becoming pregnant my child will be healthy | | | | | | | | | ☐ |

Please answer each of the following questions by moving the marker to the number that best describes your opinion. Some of the questions may appear to be similar, but they do address somewhat different issues.

| Extremely likely | | | | | Extremely unlikely | | |
|---|---|---|---|---|---|---|---|
| 0 | 1 | 2 | 3 | 4 | 5 | 6 | 7 |

| | Extremely likely 0 | 1 | 2 | 3 | 4 | 5 | Extremely unlikely 6 | 7 |
|---|---|---|---|---|---|---|---|---|
| If I take good care of my health for 6 months before becoming pregnant my child will be healthy | | | | | | | | |
| I intend to exercise regularly for at least 6 months before becoming pregnant | | | | | | | | |
| I am confident I can exercise regularly for 6 months before becoming pregnant | | | | | | | | |
| I am confident I can avoid drinking alcohol for 6 months before becoming pregnant | | | | | | | | |
| I will make an effort to exercise regularly for at least 6 months before becoming pregnant | | | | | | | | |
| I will make an effort to avoid alcohol for at least 6 months before becoming pregnant | | | | | | | | |
| I am confident I can eat a healthy diet for 6 months before becoming pregnant | | | | | | | | |
| Avoiding alcohol for 6 months before becoming pregnant is up to me | | | | | | | | |

| | Extremely likely 0 | 1 | 2 | 3 | 4 | Extremely unlikely 5 | 6 | 7 |
|---|---|---|---|---|---|---|---|---|
| I intend to avoid alcohol for at least 6 months before becoming pregnant | | | | | | | | |
| I intend to eat a heathy diet for at least 6 months before becoming pregnant | | | | | | | | |
| Exercising regularly for 6 months before becoming pregnant is up to me | | | | | | | | |
| I will make an effort to eat a heathy diet for at least 6 months before becoming pregnant | | | | | | | | |
| Eating a healthy diet for 6 months before becoming pregnant is up to me | | | | | | | | |
| If I do not smoke for 6 months before becoming pregnant my child will be healthy | | | | | | | | |

#### Please answer each of the following questions by moving the marker to the number that best describes your opinion. Some of the questions may appear to be similar, but they do address somewhat different issues.

| Strongly agree 0 | 1 | 2 | 3 | 4 | Strongly disagree 5 | 6 | 7 |
|---|---|---|---|---|---|---|---|

| | Strongly agree 0 | 1 | 2 | 3 | 4 | Strongly disagree 5 | 6 | 7 |
|---|---|---|---|---|---|---|---|---|
| My reproductive partner thinks it's important for me to exercise regularly for 6 months before becoming pregnant | | | | | | | | |
| My reproductive partner thinks it's important for me to eat a healthy diet for 6 months before becoming pregnant | | | | | | | | |
| My reproductive partner thinks it's important for me to avoid alcohol for 6 months before becoming pregnant. | | | | | | | | |

Please answer each of the following questions by moving the marker to the number that best describes your opinion. Some of the questions may appear to be similar, but they do address somewhat different issues.

| | Strongly agree 0 | 1 | 2 | 3 | 4 | Strongly disagree 5 | 6 | 7 |
|---|---|---|---|---|---|---|---|---|
| My older female relatives (e.g., mother, grandmother, aunt) think it's important for me to exercise regularly for 6 months before becoming pregnant | | | | | | | | |

| | Strongly agree 0 | 1 | 2 | 3 | 4 | 5 | 6 | Strongly disagree 7 |
|---|---|---|---|---|---|---|---|---|
| My older female relatives (e.g., mother, grandmother, aunt) think it’s important for me to eat a healthy diet for 6 months before becoming pregnant | | | | | | | | |
| My older female relatives (e.g., mother, grandmother, aunt) think it’s important for me to avoid alcohol for 6 months before becoming pregnant | | | | | | | | |

Please answer each of the following questions by moving the marker to the number that best describes your opinion. Some of the questions may appear to be similar, but they do address somewhat different issues.

| | Strongly agree 0 | 1 | 2 | 3 | 4 | 5 | 6 | Strongly disagree 7 |
|---|---|---|---|---|---|---|---|---|
| My older male relatives (e.g., father, grandfather, uncle) think it’s important for me to exercise regularly for 6 months before becoming pregnant | | | | | | | | |

| | Strongly agree | | | | | Strongly disagree | | | |
|---|---|---|---|---|---|---|---|---|---|
| | 0 | 1 | 2 | 3 | 4 | 5 | 6 | 7 | |
| My older male relatives (e.g., father, grandfather, uncle) think it’s important for me to eat a healthy diet for 6 months before becoming pregnant | | | | | | | | | ☐ |
| My older male relatives (e.g., father, grandfather, uncle) think it’s important for me to avoid alcohol for 6 months before becoming pregnant | | | | | | | | | ☐ |

Please answer each of the following questions by moving the marker to the number that best describes your opinion. Some of the questions may appear to be similar, but they do address somewhat different issues.

| | Strongly agree | | | | | Strongly disagree | | | |
|---|---|---|---|---|---|---|---|---|---|
| | 0 | 1 | 2 | 3 | 4 | 5 | 6 | 7 | |
| My male friends and family of similar age thinks it’s important for me to exercise regularly for 6 months before becoming pregnant | | | | | | | | | ☐ |

| | Strongly agree 0 | 1 | 2 | 3 | 4 | 5 | 6 | Strongly disagree 7 |
|---|---|---|---|---|---|---|---|---|
| My male friends and family of similar age thinks it's important for me to eat a healthy diet for 6 months before becoming pregnant | | | | | | | | |
| My male friends and family of similar age thinks it's important for me to avoid alcohol for 6 months before becoming pregnant . | | | | | | | | |

Please answer each of the following questions by moving the marker to the number that best describes your opinion. Some of the questions may appear to be similar, but they do address somewhat different issues.

| | Strongly agree 0 | 1 | 2 | 3 | 4 | 5 | 6 | Strongly disagree 7 |
|---|---|---|---|---|---|---|---|---|
| My female friends and family of similar age thinks it's important for me to exercise regularly for 6 months before becoming pregnant | | | | | | | | |

| | Strongly agree 0 | 1 | 2 | 3 | 4 | Strongly disagree 5 | 6 | 7 |
|---|---|---|---|---|---|---|---|---|
| My female friends and family of similar age thinks it's important for me to eat a healthy diet for 6 months before becoming pregnant | | | | | | | | |
| My female friends and family of similar age thinks it's important for me to avoid alcohol for 6 months before becoming pregnant | | | | | | | | |

Please answer each of the following questions by moving the marker to the number that best describes your opinion. Some of the questions may appear to be similar, but they do address somewhat different issues.

| | Strongly agree 0 | 1 | 2 | 3 | 4 | Strongly disagree 5 | 6 | 7 |
|---|---|---|---|---|---|---|---|---|
| My health care provider thinks it's important for me to exercise regularly for 6 months before becoming pregnant | | | | | | | | |
| My health care provider thinks it's important for me to eat a healthy diet for 6 months before becoming pregnant | | | | | | | | |

| | Strongly agree 0 | 1 | 2 | 3 | 4 | 5 | 6 | Strongly disagree 7 |
|---|---|---|---|---|---|---|---|---|
| My health care provider thinks it's important for me to avoid alcohol for 6 months before becoming pregnant | | | | | | | | |

#### Section 4: Prepregnancy health information and advice

Before you became pregnant, did you look at any information about becoming pregnant?

Yes, I went looking for information and found it

Yes, I did not go looking for information but found it anyway

No, I went looking for information but did not find any

No, I did not go looking for information and did not find any

Which sources of pre-pregnancy health and care information can you remember seeing before you became pregnant? (*Tick all that apply*)

Books

Leaflets

Internet

Magazines

Family or friends

General practitioner (GP)

Midwife

Obstetrician/Gynecologist

Pharmacist

Naturopath

Doula

Other (please give details)

#### To whom did the pre-pregnancy health and care information you saw relate to?

Me only

My partner only

Both me and my partner

#### In the year before you become pregnant did you visit a health professional about becoming pregnant?

Yes

No

#### If you were given pre-pregnancy health and care information by a health professional, who was the provided pre-pregnancy information about?

| | Health professional visited for advice | Information about... | | | |
|---|---|---|---|---|---|
| | Did not visit | Me only | My partner only | Me and my partner | None provided |
| GP | ☐ | ○ | ○ | ○ | ○ |
| Midwife | ☐ | ○ | ○ | ○ | ○ |
| Naturopath | ☐ | ○ | ○ | ○ | ○ |
| Other (please give details) [ ] | ☐ | ○ | ○ | ○ | ○ |

Before you became pregnant, did anyone give you information about any of the following topics within the context of preparing for pregnancy?

(a) If ‘No’, please tick the box

(b) If ‘Yes’, please indicate who provided the information

| | (a) | (b) Yes, the information was provided by: | | | | |
|---|---|---|---|---|---|---|
| | No | GP | Midwife | Obstetrician/ Gynecologist | Naturopath | Family/ Friends |
| Eating a healthy diet | ☐ | ☐ | ☐ | ☐ | ☐ | ☐ |
| Being the right weight for your height | ☐ | ☐ | ☐ | ☐ | ☐ | ☐ |
| Physical activity | ☐ | ☐ | ☐ | ☐ | ☐ | ☐ |
| Caffeine | ☐ | ☐ | ☐ | ☐ | ☐ | ☐ |

| | (a) | (b) Yes, the information was provided by: | | | | |
|---|---|---|---|---|---|---|
| | No | GP | Midwife | Obstetrician/ Gynecologist | Naturopath | Family/ Friends |
| Alcohol | ☐ | ☐ | ☐ | ☐ | ☐ | ☐ |
| Smoking | ☐ | ☐ | ☐ | ☐ | ☐ | ☐ |
| Street drugs | ☐ | ☐ | ☐ | ☐ | ☐ | ☐ |
| Immunisation | ☐ | ☐ | ☐ | ☐ | ☐ | ☐ |
| | No | GP | Midwife | Obstetrician/ Gynecologist | Naturopath | Family/ Friends |
| Dental check | ☐ | ☐ | ☐ | ☐ | ☐ | ☐ |
| Sexually transmitted infections | ☐ | ☐ | ☐ | ☐ | ☐ | ☐ |
| Cervical screening | ☐ | ☐ | ☐ | ☐ | ☐ | ☐ |
| Stopping contraception | ☐ | ☐ | ☐ | ☐ | ☐ | ☐ |
| Conception/fertility | ☐ | ☐ | ☐ | ☐ | ☐ | ☐ |
| Air pollution | ☐ | ☐ | ☐ | ☐ | ☐ | ☐ |
| Pesticides | ☐ | ☐ | ☐ | ☐ | ☐ | ☐ |
| Exposure to heavy metals (e.g. lead or mercury) | ☐ | ☐ | ☐ | ☐ | ☐ | ☐ |
| | No | GP | Midwife | Obstetrician/ Gynecologist | Naturopath | Family/ Friends |
| Time between pregnancies | ☐ | ☐ | ☐ | ☐ | ☐ | ☐ |
| Genetic screening | ☐ | ☐ | ☐ | ☐ | ☐ | ☐ |
| Reviewing medications | ☐ | ☐ | ☐ | ☐ | ☐ | ☐ |
| Domestic violence | ☐ | ☐ | ☐ | ☐ | ☐ | ☐ |
| Antenatal depression | ☐ | ☐ | ☐ | ☐ | ☐ | ☐ |
| Postnatal depression | ☐ | ☐ | ☐ | ☐ | ☐ | ☐ |

Some women take vitamins or supplements before they become pregnant.

<u>Before</u> you became pregnant, did anyone give you information about vitamins or supplements within the context of preparing for pregnancy?
(a) If 'No', please tick the box
(b) If 'Yes', please tick the box to indicate who provided you the information

| | (a) | (b) Yes, the information was provided by: | | | | |
|---|---|---|---|---|---|---|
| | No | GP | Midwife | Obstetrician/ Gynecologist | Naturopath | Family/ Friends |
| Folic acid | ☐ | ☐ | ☐ | ☐ | ☐ | ☐ |
| Pregnancy multivitamin | ☐ | ☐ | ☐ | ☐ | ☐ | ☐ |
| Ordinary multivitamin | ☐ | ☐ | ☐ | ☐ | ☐ | ☐ |
| Vitamin D | ☐ | ☐ | ☐ | ☐ | ☐ | ☐ |
| Iron | ☐ | ☐ | ☐ | ☐ | ☐ | ☐ |
| Omega 3 (e.g. fish oils) | ☐ | ☐ | ☐ | ☐ | ☐ | ☐ |
| | No | GP | Midwife | Obstetrician/ Gynecologist | Naturopath | Family/ Friends |
| Vitamin C | ☐ | ☐ | ☐ | ☐ | ☐ | ☐ |
| Zinc | ☐ | ☐ | ☐ | ☐ | ☐ | ☐ |
| Iodine | ☐ | ☐ | ☐ | ☐ | ☐ | ☐ |
| Calcium | ☐ | ☐ | ☐ | ☐ | ☐ | ☐ |
| Herbal medicines | ☐ | ☐ | ☐ | ☐ | ☐ | ☐ |

| | (a) | (b) Yes, the information was provided by: | | | | |
|---|---|---|---|---|---|---|
| | No | GP | Midwife | Obstetrician/ Gynecologist | Naturopath | Family/ Friends |
| Other (please give details) ☐ | ☐ | ☐ | ☐ | ☐ | ☐ | ☐ |

The next questions are similar to those asked earlier in the questionnaire. This time please answer them thinking about <u>since</u> you became pregnant with your current pregnancy.

<u>Since</u> you became pregnant, did you come across any information about becoming pregnant?

Yes, I went looking for information and found it

Yes, I did not go looking for information but found it anyway

No, I went looking for information but did not find any

No, I did not go looking for information and did not find any

Click to write the question text

Books

Leaflets

Internet

Magazines

Family or friends

Doula

General practitioner (GP)

Midwife

Pharmacist

Naturopath

Obstetrician/Gynecologist

Other (please give details)

To whom did the pre-pregnancy health and care information you saw relate to?

Me only

My partner only

Both me and my partner

Some women take vitamins or supplements before they become pregnant.

Before you became pregnant, did anyone give you information about vitamins or supplements within the context of preparing for pregnancy?
(a) If 'No', please tick the box
(b) If 'Yes', please tick the box to indicate who provided you the information

| | (a) | (b) Yes, the information was provided by: | | | | |
|---|---|---|---|---|---|---|
| | No | GP | Midwife | Obstetrician/ Gynecologist | Naturopath | Family/ Friends |
| Folic acid | ☐ | ☐ | ☐ | ☐ | ☐ | ☐ |
| Pregnancy multivitamin | ☐ | ☐ | ☐ | ☐ | ☐ | ☐ |
| Ordinary multivitamin | ☐ | ☐ | ☐ | ☐ | ☐ | ☐ |
| Vitamin D | ☐ | ☐ | ☐ | ☐ | ☐ | ☐ |
| Iron | ☐ | ☐ | ☐ | ☐ | ☐ | ☐ |
| Omega 3 (e.g. fish oils) | ☐ | ☐ | ☐ | ☐ | ☐ | ☐ |
| Vitamin C | ☐ | ☐ | ☐ | ☐ | ☐ | ☐ |
| | No | GP | Midwife | Obstetrician/ Gynecologist | Naturopath | Family/ Friends |
| Zinc | ☐ | ☐ | ☐ | ☐ | ☐ | ☐ |
| Iodine | ☐ | ☐ | ☐ | ☐ | ☐ | ☐ |
| Calcium | ☐ | ☐ | ☐ | ☐ | ☐ | ☐ |
| Herbal medicines | ☐ | ☐ | ☐ | ☐ | ☐ | ☐ |
| Other (please give details) [ ] | ☐ | ☐ | ☐ | ☐ | ☐ | ☐ |
| Other (please give details) [ ] | ☐ | ☐ | ☐ | ☐ | ☐ | ☐ |

Did anyone give you information about any of the following topics since you became pregnant?

(a) If ‘No’, please tick the box for each topic you did not receive information

(b) If ‘Yes’, please tick the box to indicate who provided the information

| | (a) | (b) Yes, the information was provided by: | | | | |
|---|---|---|---|---|---|---|
| | No | GP | Midwife | Obstetrician/ Gynecologist | Naturopath | Family/ Friends |
| Eating a healthy diet | ☐ | ☐ | ☐ | ☐ | ☐ | ☐ |
| Being the right weight for your height | ☐ | ☐ | ☐ | ☐ | ☐ | ☐ |
| Physical activity | ☐ | ☐ | ☐ | ☐ | ☐ | ☐ |
| Caffeine | ☐ | ☐ | ☐ | ☐ | ☐ | ☐ |
| Alcohol | ☐ | ☐ | ☐ | ☐ | ☐ | ☐ |
| Smoking | ☐ | ☐ | ☐ | ☐ | ☐ | ☐ |
| Street drugs | ☐ | ☐ | ☐ | ☐ | ☐ | ☐ |
| | No | GP | Midwife | Obstetrician/ Gynecologist | Naturopath | Family/ Friends |
| Immunisation | ☐ | ☐ | ☐ | ☐ | ☐ | ☐ |
| Dental check | ☐ | ☐ | ☐ | ☐ | ☐ | ☐ |
| Sexually transmitted infections | ☐ | ☐ | ☐ | ☐ | ☐ | ☐ |
| Cervical screening | ☐ | ☐ | ☐ | ☐ | ☐ | ☐ |
| Stopping contraception | ☐ | ☐ | ☐ | ☐ | ☐ | ☐ |
| Conception/fertility | ☐ | ☐ | ☐ | ☐ | ☐ | ☐ |

| | (a) | (b) Yes, the information was provided by: | | | | |
|---|---|---|---|---|---|---|
| | No | GP | Midwife | Obstetrician/ Gynecologist | Naturopath | Family/ Friends |
| Air pollution | ☐ | ☐ | ☐ | ☐ | ☐ | ☐ |
| | No | GP | Midwife | Obstetrician/ Gynecologist | Naturopath | Family/ Friends |
| Pesticides | ☐ | ☐ | ☐ | ☐ | ☐ | ☐ |
| Exposure to heavy metals (e.g. lead or mercury) | ☐ | ☐ | ☐ | ☐ | ☐ | ☐ |
| Time between pregnancies | ☐ | ☐ | ☐ | ☐ | ☐ | ☐ |
| Domestic violence | ☐ | ☐ | ☐ | ☐ | ☐ | ☐ |
| Antenatal depression | ☐ | ☐ | ☐ | ☐ | ☐ | ☐ |
| Postnatal depression | ☐ | ☐ | ☐ | ☐ | ☐ | ☐ |

#### Section 4: Your Health History

How would you describe your current health generally? Would you say it is (please tick one)

Excellent

Good

Fair

Poor

Do you have any longstanding illness, disability or infirmity? (Note: *Longstanding* means anything that has troubled your over a period of time or that is likely to affect you over a period of time)

Yes

No

In the 3 months before you became pregnant were you taking any medications/receiving treatment for OR had been diagnosed with any of the conditions listed below? (*Please tick all that apply*)

Acne

ADHD

Allergies

Anaemia

Anxiety

Asthma

Bipolar disorder

Coeliac disease

Depression

Insulin-dependent diabetes

Non insulin-dependent diabetes

Eating disorder (e.g. anorexia, bulemia)

Endometriosis

Epilepsy

Heart disease

High blood pressure

HIV

Kidney disease

Lung disease

Lupus

Polycystic ovarian syndrome (PCOS)

Phenylketonuria (PKU)

Sexually transmitted infection

Other (please give details)

How many pregnancies have you had before this current pregnancy? (Please put the number. If none, put ‘0’)

Have you had any pregnancies that were...(Please put the number. If none, put ’0’)

Miscarriage

Ectopic pregnancy (or tubal pregnancy)

Still birth (i.e. birth of a baby that is not alive, past 24 weeks pregnancy)

Terminated because of fetal/genetic abnormalities

Other (please give details)

*If you have experienced any of the above conditions you might like to someone about how you are feeling. You could ring Lifeline on 131114 (local call)*

#### Have you experienced any of the following symptoms or conditions in previous pregnancies? (Please tick all that apply)

Gestational diabetes

Pre-eclampsia

Repeated vomiting

High blood pressure (hypertension)

Vaginal bleeding

#### Have you given birth to any children?

Yes

No

#### What is the year of birth for each of your children?

Child 1

Child 2

Child 3

Child 4

Child 5

Child 6

Please complete the table below for each child. Please tick if they have had any of the medical conditions. (Please tick all that apply)

| | Child 1 | Child 2 | Child 3 | Child 4 | Child 5 | Child 6 |
|---|---|---|---|---|---|---|

| | Child 1 | Child 2 | Child 3 | Child 4 | Child 5 | Child 6 |
|---|---|---|---|---|---|---|
| | Yes | Yes | Yes | Yes | Yes | Yes |
| Born more than 3 weeks prematurely | ☐ | ☐ | ☐ | ☐ | ☐ | ☐ |
| Low birth weight (less than 2.5kg/5lb) | ☐ | ☐ | ☐ | ☐ | ☐ | ☐ |
| Allergies | ☐ | ☐ | ☐ | ☐ | ☐ | ☐ |
| Asthma | ☐ | ☐ | ☐ | ☐ | ☐ | ☐ |
| Autism | ☐ | ☐ | ☐ | ☐ | ☐ | ☐ |
| Cerebral palsy | ☐ | ☐ | ☐ | ☐ | ☐ | ☐ |
| Cleft lip or palate | ☐ | ☐ | ☐ | ☐ | ☐ | ☐ |
| Downs syndrome | ☐ | ☐ | ☐ | ☐ | ☐ | ☐ |
| Neural tube defects | ☐ | ☐ | ☐ | ☐ | ☐ | ☐ |
| Spina bifida | ☐ | ☐ | ☐ | ☐ | ☐ | ☐ |
| Other major health condition or illness | ☐ | ☐ | ☐ | ☐ | ☐ | ☐ |

#### Block 7

What is today's date?

Have we missed anything? If you have anything else you would like to tell us, please write it below

Powered by Qualtrics
