## Supplementary File 2 for "WOMEN’S PRECONCEPTION HEALTH, PLANNING, AND BEHAVIOURS: A CROSS-SECTIONAL SURVEY OF PREGNANT WOMEN IN AUSTRALIA"

Supplementary File 2: London Measure of Unplanned Pregnancy Item Responses and scores*

| In the month that I became pregnant… (n=613) | n (%) |
| --- | --- |
| *Not using contraception* | 552 (90.1) |
| *Using contraception, but inconsistent* | 39 (6.4) |
| *Always used contraception but knew that the method had failed at least once* | 8 (1.3) |
| *Always used contraception* | 14 (2.3) |
| Felt pregnancy occurred at the… (n=613) |  |
| *Right time* | 451 (73.6) |
| *Ok, but not quite right time* | 146 (23.8) |
| *Wrong time* | 16 (2.6) |
| Just before I became pregnant… (n=613) |  |
| *I intended to get pregnant* | 454 (74.1) |
| *My intentions kept changing* | 76 (12.4) |
| *I did not intend to get pregnant* | 83 (13.5) |
| Just before I became pregnant… (n=613) |  |
| *I wanted to have a baby* | 471 (76.8) |
| *I had mixed feelings about having a baby* | 131 (21.4) |
| *I did not want to have a baby* | 11 (1.8) |
| Before I became pregnant… (n=611) |  |
| *My partner and I had agreed that we would like me to become pregnant* | 491 (80.4) |
| *My partner and I had discussed having children together, but hadn’t agreed for me to get pregnant* | 105 (17.2) |
| *We never discussed having children together* | 8 (1.3) |
| *I chose to become pregnant without a partner* | 7 (1.2) |
| Before you became pregnant, did you do anything to improve your health in preparation for your pregnancy? (n=549)* |  |
| *Took folic acid* | 153 (27.8) |
| *Took iodine* | 2 (0.3) |
| *Took Vitamin D* | 5 (0.9) |
| *Stopped or cut down smoking* | 17 (3.0) |
| *Stopped or cut down drinking alcohol* | 25 (4.5) |
| *Ate more healthily* | 37 (6.7) |
| *Sought medical/health advice* | 65 (11.8) |
| *Took some other action* | 85 (15.5) |
| *I did not do any of the above before my pregnancy* | 160 (29.1) |
| Level of Pregnancy Planning Score |  |
| *Planned (10-11)* | 364 (59.6) |
| *Ambivalent (4-9)* | 234 (38.3) |
| *Unplanned (0-3)* | 13 (2.1) |

*****This item was set up to allow only one response option rather than multiple response options and as such was not scored
